# Acceptability, Safety and Feasibility of a Sedentary Behaviour Intervention (REACH-SCI) in Individuals with Spinal Cord Injury: A Mixed-Methods Study

**DOI:** 10.64898/2026.10.21.26365989

**Authors:** Daniel L. Cooper, Alyson Warland, Emma Norris, Cherry Kilbride, Sue Paddison, Benjamin D. Maylor, Katherine A. Finlay, Louise Ferrandino, Daniel P. Bailey

**Author notes:** Corresponding author: Daniel L Cooper.

## Abstract

**Purpose:** Individuals with paraplegia often have high levels of sedentary behaviour, which increases cardiovascular disease risk. This study assessed the acceptability, safety and feasibility of a sedentary behaviour intervention (REACH-SCI) in individuals with paraplegia.

**Materials and methods:** A pre-post feasibility study was conducted in wheelchair-users with paraplegia (n=18) to evaluate an 8-week intervention comprised of a wearable activity tracker, educational booklet, goal setting, motivational support sessions, peer support and activity tools (Trial registration: NCT06957483). Acceptability was assessed via semi-structured interviews and questionnaires, safety via adverse events, pain and fatigue, and intervention feasibility via components being delivered as intended. Intervention engagement and preliminary efficacy were also assessed.

**Results:** The intervention and study procedures were acceptable and safe, intervention components were delivered as planned, and the study was feasible in terms of participant recruitment, retention and data completion rates. Engagement with intervention components was generally high, except for peer support. The intervention demonstrated preliminary efficacy for improving sedentary behaviour, physical activity, cardiovascular disease biomarkers and psychosocial outcomes.

**Conclusions:** An intervention to reduce sedentary behaviour in individuals with paraplegia was acceptable, safe and feasible. The effectiveness of REACH-SCI should be evaluated in a definitive trial to inform clinical care and public health guidelines.

## Introduction

Spinal cord injury (SCI) is associated with a number of physiological changes related to body composition and metabolic function that increase cardiovascular and metabolic risk [1–4]. Consequently, individuals with SCI have an elevated risk of cardiovascular disease (CVD), with the likelihood of heart disease and stroke estimated to be 2.72- and 3.72-times higher, respectively, than the general population [5]. Therefore, interventions targeting modifiable CVD risk factors are urgently needed in individuals with SCI.

Sedentary behaviour, defined as an energy expenditure of < 1.5 metabolic equivalents while sitting, lying or reclined during waking hours [6], is an independent risk factor for CVD in the general population [7,8]. Postma et al. [9] reported that individuals with SCI were sedentary for 12.2 hours/day, compared to 9.6 hours/day in the general population [10], which may explain the higher CVD risk in this population. Paraplegia, defined as a SCI at or below the first thoracic vertebrae, results in lower body paralysis whilst preserving upper limb function [11]. Consequently, individuals with paraplegia have an opportunity to break up sedentary behaviour through upper limb movement. Reducing and breaking up sedentary behaviour could, therefore, potentially represent modifiable targets for lowering CVD risk in this population.

Whilst sedentary behaviour interventions are acceptable, safe and feasible in the general population [12–14], interventions developed for ambulatory individuals may not be directly transferable to wheelchair-users with paraplegia. This population experiences distinct barriers to reducing sedentary behaviour, including loss of movement in the lower limbs and trunk, neuropathic pain and reliance on wheelchairs that can be difficult to propel [15]. Following discharge into the community these barriers may be exacerbated, which could explain the corresponding decrease in activity levels observed after individuals with SCI return to their home environment [16]. Therefore, interventions targeting sedentary behaviour are needed to support individuals at the early stages of the SCI care pathway after inpatient discharge, as well as those with a long-term injury. However, there are a lack of interventions targeting sedentary behaviour in individuals with paraplegia [17]. To address this gap, a novel intervention, REACH-SCI (Reducing sedEntary Activities to improve Cardiovascular Health in individuals with Spinal Cord Injury), was co-designed with key stakeholders tailored specifically to wheelchair-users with paraplegia [15].

The aim of this study was to assess the acceptability, safety and feasibility of the REACH-SCI intervention in individuals with paraplegia at different stages of SCI rehabilitation. The primary objectives were to explore intervention acceptability using an integrated mixed-methods approach, including the extent to which qualitative findings explained, confirmed, and/or expanded on quantitative data, as well as safety and feasibility. Secondary objectives were to evaluate participant engagement with the intervention, the acceptability of data collection procedures, the feasibility of recruiting and retaining participants, and preliminary efficacy for improving sedentary behaviour, physical activity, CVD biomarkers and psychosocial outcomes.

## Methods

### Study Overview

This study used a mixed-methods, single-arm, pre-post design (see study schematic in Figure 1). Measurements were taken at baseline and repeated immediately following the eight-week intervention. Semi-structured interviews were undertaken with all participants post-intervention. Ethical approval was granted from Brunel University of London College of Health, Medicine and Life Sciences Research Ethics Committee (50038-NHS-Mar/2025-54084-2) and the National Health Service East of England - Cambridge South Research Ethics Committee (25/EE/0090). The study adhered to the principles of the Declaration of Helsinki. Consolidated Standards of Reporting Trials guidelines for feasibility and pilot trials [18] and Good Reporting of a Mixed-Methods Study (GRAMMS) guidelines [19] are followed for reporting purposes. The study protocol was prospectively registered with clinicaltrials.gov (NCT06957483).

**Figure 1.**
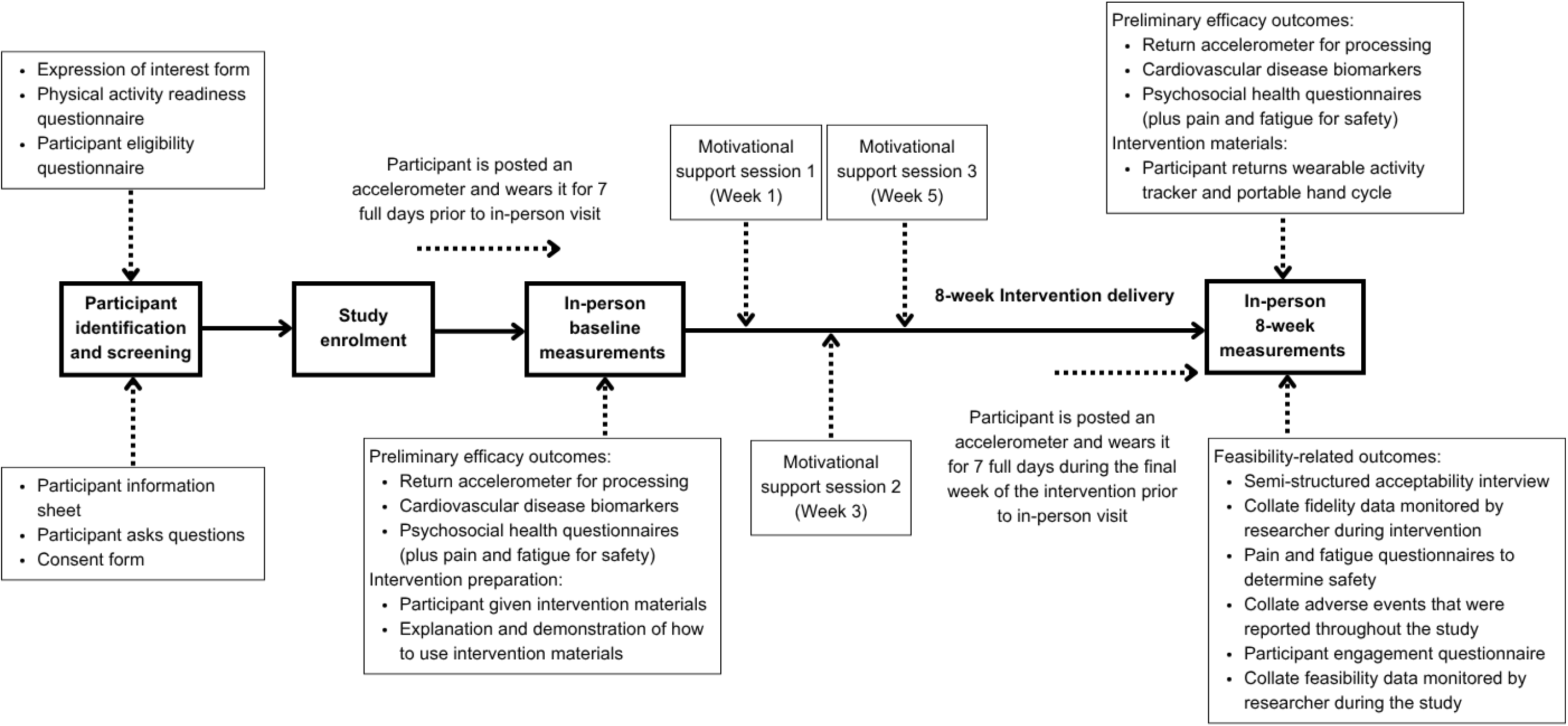
Study schematic.

### Study Setting

The intervention was delivered remotely, increasing accessibility to the study, to community-dwelling individuals with paraplegia. Outcome measurements and semi-structured interviews were conducted within a laboratory and private interview room, respectively, in the Department of Sport, Health and Exercise Sciences at Brunel University of London.

### Participant Eligibility Criteria

Participant inclusion criteria were adults (≥ 18 years) with a self-reported SCI at T1 or below [11], irrespective of lesion completeness or cause, who used a manual wheelchair as their primary mode of mobility inside and outside of the home. Participants needed to be able to travel to Brunel University of London, have regular access to a smartphone or tablet with reliable internet connection, and communicate in English at a level sufficient to participate in the study. Additionally, participants needed to be able to provide informed consent, provide fasted fingertip blood samples during measurement sessions, and independently transfer to a treatment couch and lay supine for anthropometric and body composition measurements.

Exclusion criteria were self-reported heart conditions, chest pain, dizziness, bone or joint problem, or any other condition that might be exacerbated by physical activity as indicated by responses to the Physical Activity Readiness Questionnaire Plus [20]. Individuals were also excluded if they had experienced uncontrolled autonomic dysreflexia in the past 12 months, were pregnant, or were participating in another study which could influence the present intervention measurements.

### Participant Recruitment

The study aimed to include participants from across the SCI care pathway from discharge from inpatient rehabilitation to more than one year post-discharge. Recruitment was undertaken through community and clinical routes including disability sport and SCI support pages on social media (Facebook and X), SCI charities (Spinal Injuries Association and Spinal Research), snowballing and previous study participants. Participants were also recruited from the London Spinal Cord Injury Centre, Royal National Orthopaedic Hospital NHS Trust during routine appointments, inpatient care, mail-out and display of posters and leaflets.

### Sample Size

The target sample size (n = 20) was guided by the primary study aims regarding intervention acceptability, safety and feasibility [21]. A power calculation was not undertaken in line with recommendations for feasibility studies [21].

### The REACH-SCI Intervention

As reported previously [15], the REACH-SCI intervention (see Table 1) was developed in accordance with the Medical Research Council framework for developing complex interventions [22] and co-designed with key stakeholders using the Behaviour Change Wheel [23]. The multi-component intervention was delivered for eight weeks following baseline measures. Intervention content is reported following the Template for Intervention Description and Replication checklist [24].

**Table 1.**
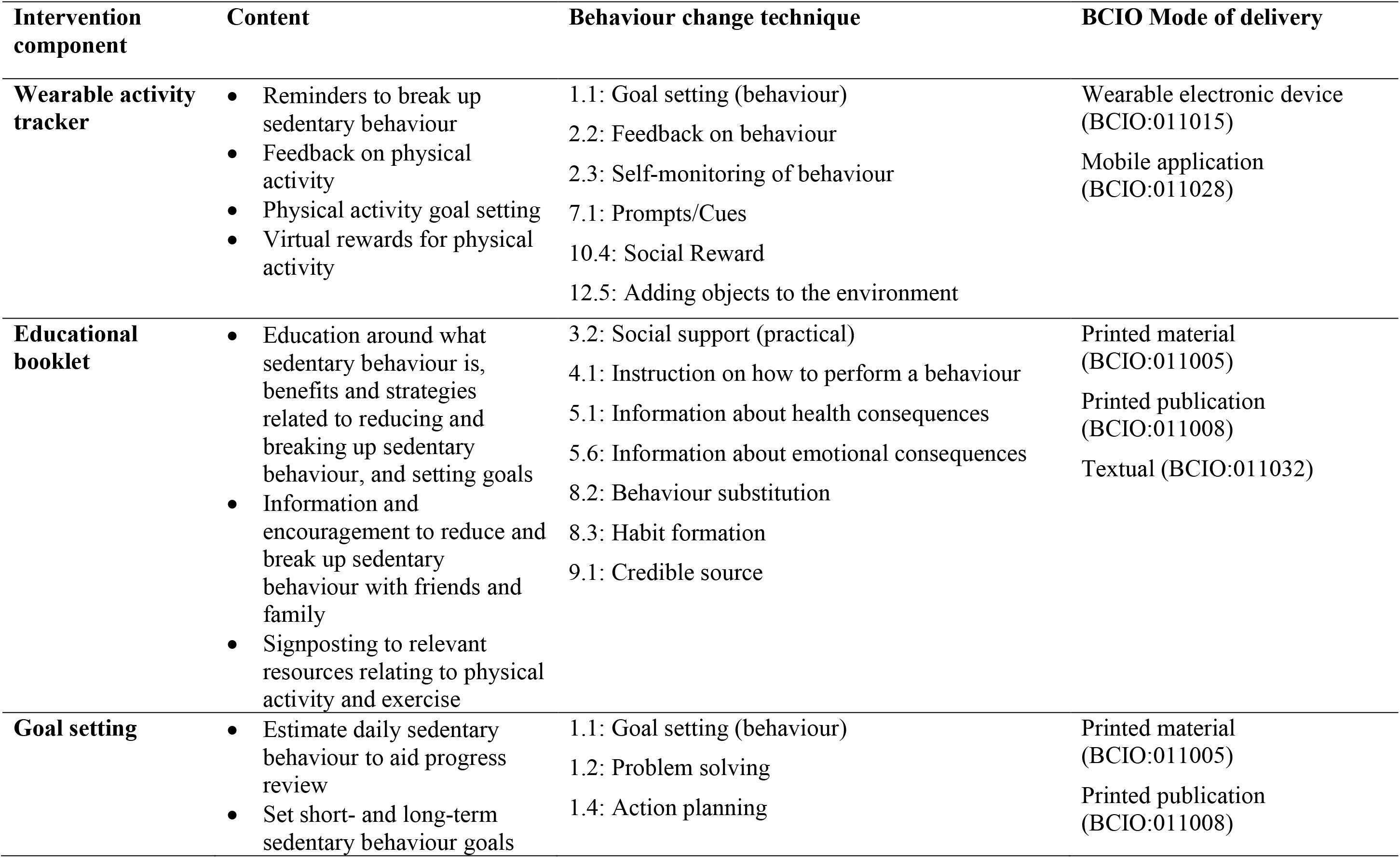
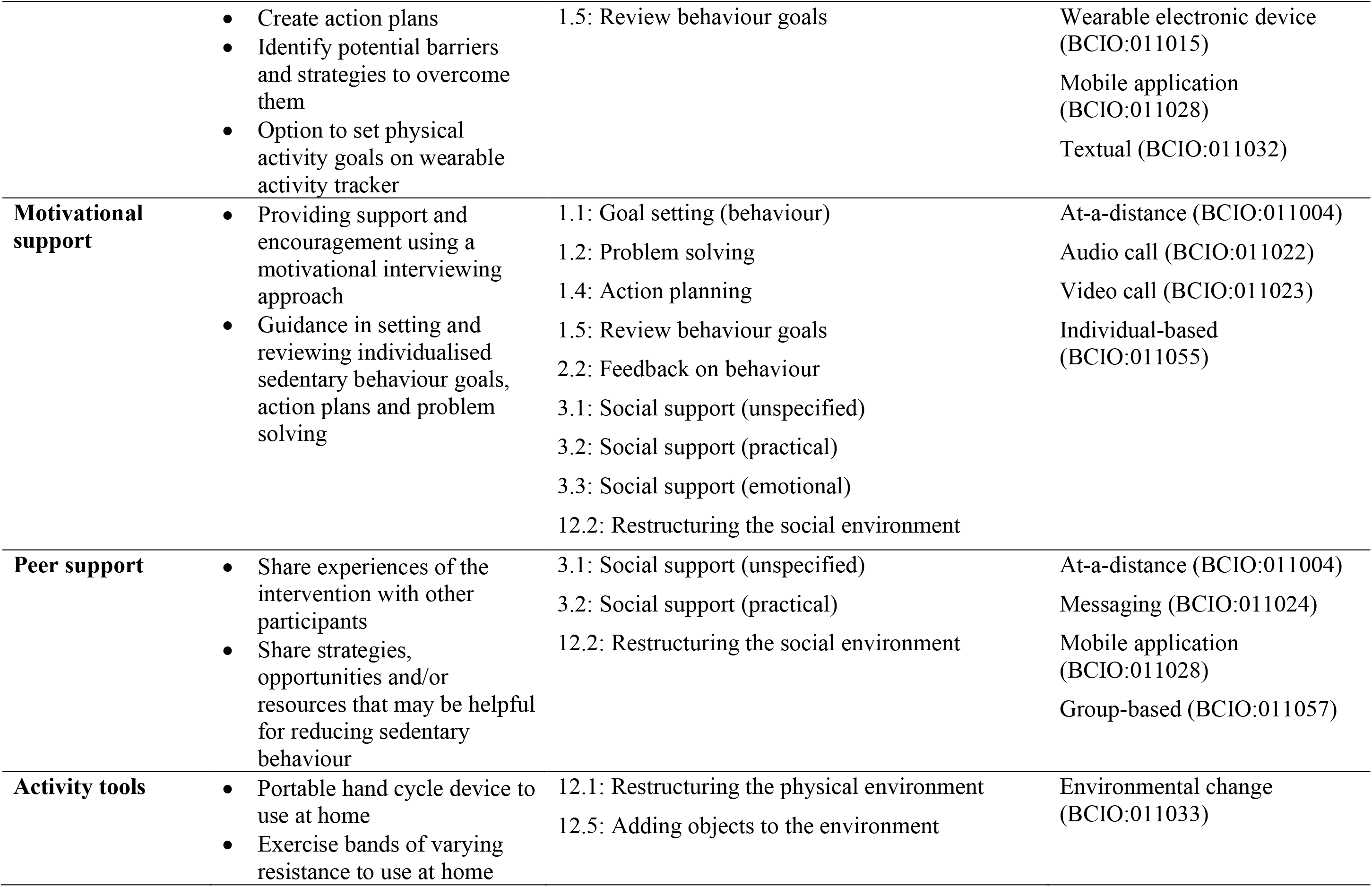
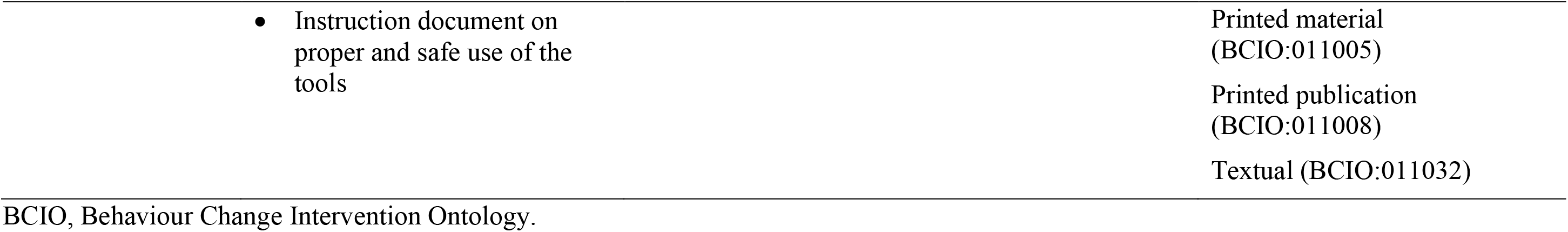
Intervention component content, behaviour change techniques and delivery mode.

| **Intervention component** | **Content** | **Behaviour change technique** | **BCIO Mode of delivery** |
| --- | --- | --- | --- |
| **Wearable activity tracker** | Reminders to break up sedentary behaviour<br>Feedback on physical activity<br>Physical activity goal setting<br>Virtual rewards for physical activity | 1.1: Goal setting (behaviour)<br>2.2: Feedback on behaviour<br>2.3: Self-monitoring of behaviour<br>7.1: Prompts/Cues<br>10.4: Social Reward<br>12.5: Adding objects to the environment | Wearable electronic device (BCIO:011015)<br>Mobile application (BCIO:011028) |
| **Educational booklet** | Education around what sedentary behaviour is, benefits and strategies related to reducing and breaking up sedentary behaviour, and setting goals<br>Information and encouragement to reduce and break up sedentary behaviour with friends and family<br>Signposting to relevant resources relating to physical activity and exercise | 3.2: Social support (practical)<br>4.1: Instruction on how to perform a behaviour<br>5.1: Information about health consequences<br>5.6: Information about emotional consequences<br>8.2: Behaviour substitution<br>8.3: Habit formation<br>9.1: Credible source | Printed material (BCIO:011005)<br>Printed publication (BCIO:011008)<br>Textual (BCIO:011032) |
| **Goal setting** | Estimate daily sedentary behaviour to aid progress review<br>Set short- and long-term sedentary behaviour goals | 1.1: Goal setting (behaviour)<br>1.2: Problem solving<br>1.4: Action planning | Printed material (BCIO:011005)<br>Printed publication (BCIO:011008) |
|  | • Create action plans<br>• Identify potential barriers and strategies to overcome them<br>• Option to set physical activity goals on wearable activity tracker | 1.5: Review behaviour goals | Wearable electronic device (BCIO:011015)<br>Mobile application (BCIO:011028)<br>Textual (BCIO:011032) |
| **Motivational support** | • Providing support and encouragement using a motivational interviewing approach<br>• Guidance in setting and reviewing individualised sedentary behaviour goals, action plans and problem solving | 1.1: Goal setting (behaviour)<br>1.2: Problem solving<br>1.4: Action planning<br>1.5: Review behaviour goals<br>2.2: Feedback on behaviour<br>3.1: Social support (unspecified)<br>3.2: Social support (practical)<br>3.3: Social support (emotional)<br>12.2: Restructuring the social environment | At-a-distance (BCIO:011004)<br>Audio call (BCIO:011022)<br>Video call (BCIO:011023)<br>Individual-based (BCIO:011055) |
| **Peer support** | • Share experiences of the intervention with other participants<br>• Share strategies, opportunities and/or resources that may be helpful for reducing sedentary behaviour | 3.1: Social support (unspecified)<br>3.2: Social support (practical)<br>12.2: Restructuring the social environment | At-a-distance (BCIO:011004)<br>Messaging (BCIO:011024)<br>Mobile application (BCIO:011028)<br>Group-based (BCIO:011057) |
| **Activity tools** | • Portable hand cycle device to use at home<br>• Exercise bands of varying resistance to use at home | 12.1: Restructuring the physical environment<br>12.5: Adding objects to the environment | Environmental change (BCIO:011033) |

BCIO, Behaviour Change Intervention Ontology.

#### Wearable Activity Tracker

Participants were provided with a Garmin Vivoactive 5 smartwatch (Garmin, Olathe, KS, USA), which was set to Wheelchair Mode and connected to the participants’ Garmin Connect smartphone app. Verbal guidance and demonstration of how to use the device were provided by the researcher during the baseline visit, following study measurements. The activity tracker provided hourly prompts to break up sedentary behaviour with a haptic alert and on-screen notification. Feedback on the number of wheelchair pushes, heart rate, calories burned and minutes spent in moderate-vigorous physical activity (MVPA) is also provided. Virtual badges are awarded for achievement of personal activity goals. The number of consecutive days in which a personal push goal is achieved is recorded and displayed. Participants had the opportunity to use their own activity tracker if all planned behaviour change techniques (BCTs) could be delivered by that device.

#### Educational Booklet

Participants were provided with a printed educational booklet, which was also emailed as an electronic version, that contained information on: (1) the definition of sedentary behaviour [6], (2) the health benefits of reducing and breaking up sedentary behaviour, (3) practical strategies to support reducing and breaking up sedentary behaviour, (4) how to set and review sedentary behaviour goals, and (5) a list of different charities, organisations and websites that provide equipment, information and/or opportunities relating to physical activity and exercise. Sedentary behaviour was referred to as ‘inactivity’ on all patient-facing materials as informed by the intervention co-design process [15].

#### Goal Setting

Participants set short- and long-term goals related to sedentary behaviour and physical activity using a goal setting worksheet in printed and electronic version. Guidance from a trained researcher was provided in motivational support sessions and described in the following section. Goals were individualised, reflecting the amount and frequency at which participants aimed to reduce or break up sedentary behaviour and, optionally, increase physical activity. Participants were prompted and guided by the researcher to record all sedentary behaviour bouts during a single day in the first and final week of the intervention and calculate their total daily sedentary time at each timepoint to assess change and aid progress review. Participants were also encouraged to set physical activity goals (e.g. daily wheelchair pushes, MVPA) within the activity tracker to support reductions in sedentary behaviour.

#### Motivational Support Sessions

A researcher trained in motivational interviewing (through an accredited course by the Association for Psychological Therapies) supported participants in making behavioural changes aimed at reducing and breaking up sedentary behaviour. This included support with setting and reviewing individualised sedentary behaviour and physical activity goals, monitoring progress, and creating action plans. The one-to-one support sessions were delivered remotely via online video call or telephone depending on participant preference. Three support sessions took place at approximately weeks one, three and five of the intervention.

#### Peer Support

An optional peer-support group was set up by a researcher using the group chat function in WhatsApp. Participants were given the option to join the group following baseline measurements. Participants who opted into this component were given verbal and textual (via email) guidance on its use by a researcher and encouraged to provide peer support and advice relating to sedentary behaviour and physical activity, in addition to sharing experiences with the intervention. The researcher did not monitor the group but accessed the group once weekly to post reminder messages encouraging use by participants.

#### Activity Tools

Participants were provided with exercise bands of varying resistance (2.3, 4.5, and 6.8 kg) (HPYGN, Shenzhen, China) and a portable hand cycle (AGM, Shenzhen, China) to use at home. Verbal guidance and demonstration of how to use the activity tools were provided by the researcher during the baseline visit, following study measurements. Printed guidance detailed how and when to use the bands and hand cycle including short, frequent bouts spread across the day. Information on safety precautions for maintaining good shoulder health were provided.

### Data Collection

The schedule of study measurements is outlined in Table 2.

**Table 2.** Study measurements at each timepoint.

| Study measure | Measurement method | Timepoint |  |  |
| --- | --- | --- | --- | --- |
|  |  | Baseline | Collected throughout intervention period | 8 weeks (post-intervention) |
| Participant characteristics | Self-report questionnaire | ✓ |  |  |
| Intervention acceptability | Semi-structured interview, intervention acceptability questionnaire |  |  | ✓ |
| Intervention feasibility | Intervention component delivery checklist |  | ✓ |  |
| Safety (adverse events) | Ad-hoc self-report |  | ✓ |  |
| Safety (pain and fatigue) | MPQ and MFIS questionnaires | ✓ |  | ✓ |
| Intervention engagement | Intervention engagement questionnaire |  |  | ✓ |
| Acceptability of data collection procedures | Semi-structured interview |  |  | ✓ |
| Study feasibility | Recruitment, retention and data completion rates | ✓ |  | ✓ |
| Sedentary behaviour and physical activity | ActiGraph GT9X Link accelerometer | ✓ |  | ✓ |
| CVD biomarkers | Automated blood pressure monitor, anthropometric measuring tape, wheelchair scales, bioelectric impedance, CardioChek PA analyser (capillary blood sample) | ✓ |  | ✓ |
| Psychosocial outcomes | GAD-7, PHQ-9, ONS-4 and SF-36ww questionnaires | ✓ |  | ✓ |

CVD, cardiovascular disease; GAD-7, Generalized Anxiety Disorder 7-item; MFIS, Modified Fatigue Impact Scale; MPQ, McGill Pain Questionnaire; ONS-4, Office for National Statistics 4-item; PHQ-9, Patient Health Questionnaire 9-item; SF-36ww, Short Form 36-item walk-wheel

#### Demographic Data

At baseline, demographic information was collected via an online questionnaire (Jisc Online Surveys).

#### Intervention Acceptability

Acceptability was explored post-intervention using a questionnaire comprising 5-point Likert scales [25] and individual semi-structured interviews (35 ± 14 min; Supplementary File 1). The data collection instruments were informed by the Theoretical Framework of Acceptability (TFA): affective attitude (how an individual feels about the intervention), burden (perceived effort required to participate in the intervention), ethicality (how the intervention fits with the individual’s value system), intervention coherence (how the individual understands the intervention and how it works), opportunity costs (how the individual must give up other benefits, value or profits to engage with the intervention), perceived effectiveness (how the individual perceives the intervention as likely to achieve its purpose), and self-efficacy (the individual’s confidence that they can perform the behaviour required to participate in the intervention) [26]. The perceived effectiveness TFA construct was used to explore the acceptability of each intervention component.

#### Intervention Safety

Intervention safety was evaluated through the recording of adverse events experienced during the study. Adverse events were self-reported by participants when prompted at each contact point with a researcher and could also be reported ad-hoc throughout the study. Safety was further assessed via self-report pain using the McGill Pain Questionnaire (MPQ) [27] and fatigue, measured with the Modified Fatigue Impact Scale (MFIS) [28], both of which are validated in individuals with SCI [29,30].

#### Intervention Feasibility

Intervention feasibility was assessed through each of the intervention components being delivered as planned by the researcher, including provision of equipment and resources, plus participant attendance and duration of motivational support sessions.

#### Intervention Engagement

Engagement with each intervention component was assessed using an online questionnaire. Participants were also able to provide open-text responses to contextualise and/or elaborate on quantitative responses for each intervention component.

#### Acceptability of Data Collection Procedures

The acceptability of the data collection procedures was assessed during the semi-structured interviews exploring participants’ experiences of the study measurements.

#### Study Feasibility

Study feasibility was determined through participant recruitment and retention rates, and the proportion of missing data for sedentary time (proposed primary outcome for a definitive trial) and secondary outcomes.

#### Sedentary Behaviour and Physical Activity

Participants wore an ActiGraph GT9X Link accelerometer (ActiGraph LLC, Pensacola, FL, USA) on their non-dominant wrist for seven full, consecutive days at baseline and during week 8 of the intervention. The ActiGraph GT9X was used to provide estimates of sedentary time (min/day), number of breaks in sedentary behaviour, percentage of day in sedentary behaviour, number of and time (min/day) in prolonged sedentary bouts (continuous periods of sedentary behaviour lasting at least 30 and 60 minutes), light physical activity (min/day), MVPA (min/day) and total physical activity (light physical activity + MVPA; min/day). Participants were asked to record any periods of non-wear in a diary.

Raw tri-axial accelerometry data (*g*) across *x*, *y* and *z* axes at a sampling rate of 30 Hz were downloaded using ActiLife v6.13.4 (ActiGraph LLC, Pensacola, FL, USA) and raw .gt3x files were processed using open-source R package GGIR (v3.1-5; www.github.com/wadpac/GGIR) [31]. Criteria for valid accelerometry data were ≥ 10 hours of waking wear time on ≥ four days [32]. Data were processed in GGIR via the inbuilt functions for sensor autocalibration [33]. Raw signal data were converted into Euclidian Norm Minus One (ENMO) values with truncated negative values. These values were then aggregated into 5 second epochs and categorised as time spent in different intensities according to published cut-points for manual wheelchair-users with SCI (≤ 37 *g*/min for sedentary behaviour; > 37 to < 222 *g*/min for light physical activity; ≥ 222 *g*/min for MVPA) [34].

#### Cardiovascular Disease Biomarkers

Body composition, blood pressure and blood biomarker were assessed at baseline and post-intervention (8 ± 9 days following intervention end date) following an overnight fast. Participants were asked to abstain from exercise for 24 hours prior to assessment visits.

##### Anthropometry and Body Composition

Body mass was calculated by measuring the mass of the participant in their wheelchair (Marsden M-610 double-beam wheelchair scales; Marsden, Rotherham, UK) and subtracting the mass of the wheelchair alone. Supine height and waist circumference were measured to the nearest 0.1 cm using an anatomical tape measure. Body mass index was calculated as: kg / m^2^. Body fat mass and percentage body fat were estimated using bioelectrical impedance analysis (Bodystat 1500; Bodystat Ltd, Isle of Man, UK).

##### Blood Pressure

After five minutes of rest, blood pressure was measured using an automated blood pressure monitor (A&D UA-611, Tokyo, Japan) whilst the participant sat upright in their wheelchair. Three measurements were taken at one-minute intervals, and the average of the two lowest readings was recorded. Mean arterial pressure was calculated as:

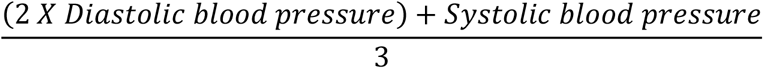

##### Blood Biomarkers

Fasting blood glucose, total cholesterol, and high-density lipoprotein (HDL) cholesterol were measured from finger prick capillary blood samples using an automated analyser (CardioChek PA; PTS Diagnostics, Indianapolis, IN, USA). Total cholesterol:HDL cholesterol ratio and non-HDL cholesterol were subsequently calculated.

#### Psychosocial Outcomes

Subjective well-being was measured using the Office for National Statistics four-item wellbeing scale [35]. Anxiety and depression were assessed using the Generalized Anxiety Disorder (GAD-7) questionnaire [36] and Patient Health Questionnaire (PHQ-9) [37], respectively. Health-related quality of life was measured using the SF-36 walk wheel, a modified version of the 36-item Short-Form Health Survey [38] validated for use in individuals with SCI [39].

### Data Analysis

#### Quantitative Data

Questionnaire data relating to intervention acceptability were analysed descriptively. Intervention feasibility was measured by calculating the rate at which each intervention component was delivered as planned. Trial and intervention safety were assessed through the number and type of adverse events reported during the study and descriptive changes in pain and fatigue.

Self-reported questionnaire data related to intervention engagement for each intervention component were analysed descriptively. Trial feasibility was assessed by calculating eligibility, recruitment, retention and study measurement completion rates. Pre-post changes in sedentary behaviour, physical activity, CVD biomarkers and psychosocial outcomes were explored using descriptive statistics (mean ± SD).

#### Qualitative Data

Semi-structured interviews were transcribed using Microsoft Teams (Microsoft Corporation, Redmond, WA, USA) and checked for accuracy by a researcher. Intervention acceptability data were analysed deductively using the TFA constructs a priori within Framework Analysis [40]. Additional inductive coding was undertaken to identify insights not captured by the TFA. Acceptability of data collection procedures were coded inductively using Framework Analysis. NVivo 12 software (Lumivero, Denver, CO, USA) was used to support data management and organisation.

#### Mixed-Methods Data Integration

Using a convergent mixed-methods approach, quantitative and qualitative data were integrated through side-by-side comparison and presented in joint display tables [41].

## Results

Eighteen participants were recruited into the study between April 2025 and September 2025. Table 3 presents the baseline characteristics of the included participants, while Figure 2 shows participant flow.

**Figure 2.**
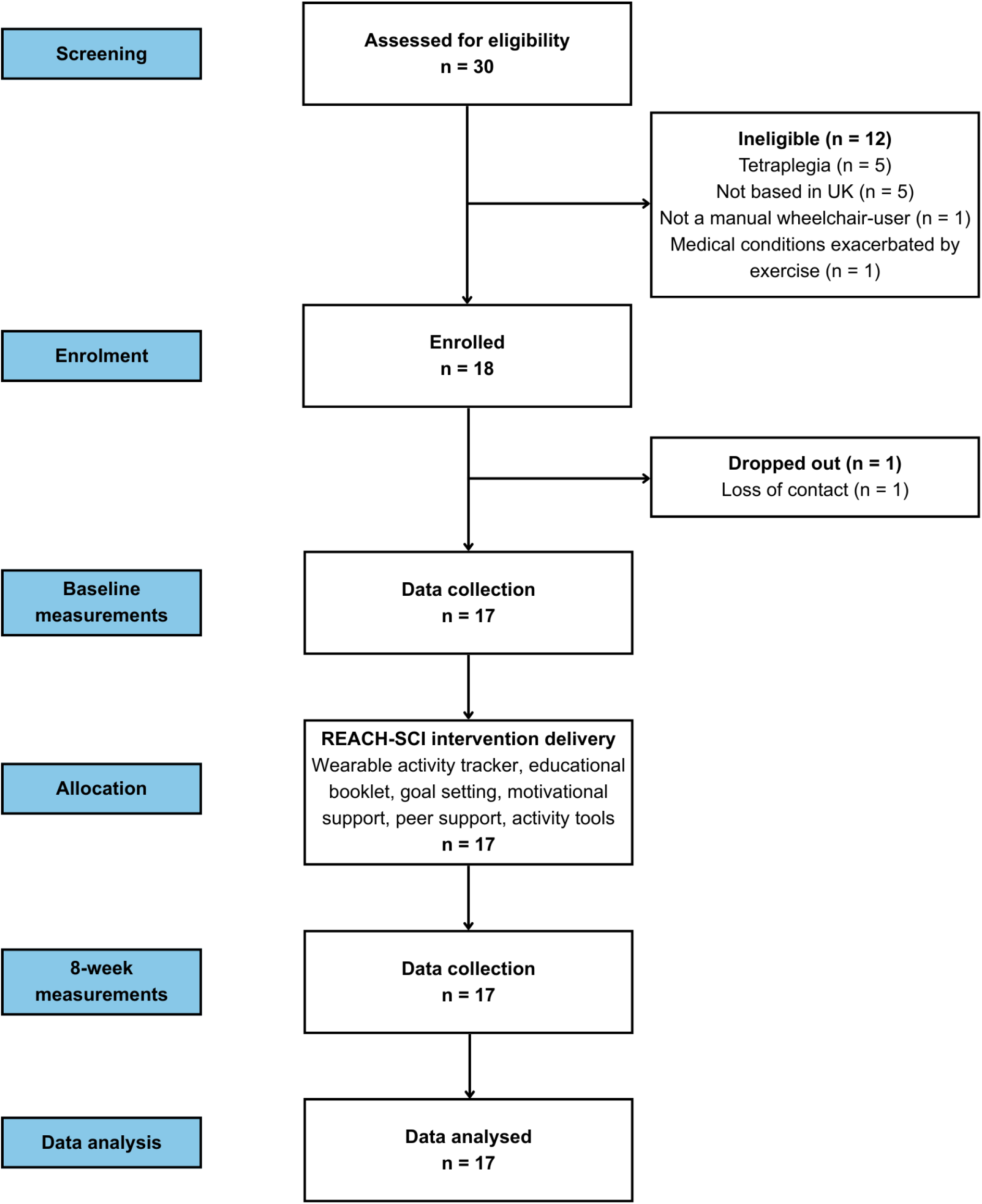
Participant flow diagram

**Table 3.** Participant characteristics (n = 18).

| Characteristic |  |  | Mean ± SD or n (%) |
| --- | --- | --- | --- |
| Sex | Female |  | 9 (50%) |
|  | Male |  | 9 (50%) |
| Ethnicity | Black British |  | 1 (6%) |
|  | White British |  | 16 (88%) |
|  | White European |  | 1 (6%) |
| Age |  |  | 51 ± 11 years |
| Time since injury |  |  | 18 ± 13 years |
| Lesion level | Thoracic | Total | 15 (83%) |
|  |  | T3 | 3 |
|  |  | T4 | 1 |
|  |  | T4 | 1 |
|  |  | T5 | 2 |
|  |  | T8 | 2 |
|  |  | T9 | 3 |
|  |  | T10 | 1 |
|  |  | T11 | 1 |
|  |  | T12 | 1 |
|  | Lumbar | Total | 3 (17%) |
|  |  | L1 | 3 |
| Completeness | Complete |  | 9 (50%) |
|  | Incomplete |  | 9 (50%) |
| Stage of SCI rehabilitation | < 1 year since initial discharge |  | 2 (11%) |
|  | ≥ 1 year since initial discharge |  | 16 (89%) |

L, lumbar; SCI, spinal cord injury; T, thoracic.

### Intervention Acceptability

#### Overall Intervention Acceptability

##### TFA Domain 1: Affective Attitude

Quantitative findings indicated high overall acceptability, with 94% of participants reporting they liked or strongly liked the intervention. Qualitative findings likewise supported this, with participants describing the intervention as enjoyable. Participants explained they liked being made aware of inactivity and that the intervention was a good challenge (Table 4; Supplementary Table 1). However, some participants felt there was too much paperwork involved in the intervention.

**Table 4.** Integrated mixed-methods intervention acceptability findings.

| **TFA domain** | **Quantitative acceptability questionnaire** | **Qualitative illustrative quotes from semi-structured interviews** | **Agreement between quantitative and qualitative data** |
| --- | --- | --- | --- |
| Affective attitude | *Did you like or dislike REACH-SCI?*<br>0% strongly dislike<br>0% dislike<br>6% no opinion<br>59% like<br>35% strongly like | “Yeah, really good... So I didn't get bored. It makes me want to carry it on even after the eight weeks” (Female, L1 Incomplete)<br>“Yes, yes [it was enjoyable]... I think being more aware of my levels of activity and kind of forcing myself to take accountability on it [was my favourite part of REACH-SCI]” (Female, T3 Incomplete)<br>“Yes [I enjoyed doing the intervention]. The challenge was good and pushing yourself to reach those goals” (Female, T5 Complete)<br>“It was like, “here’s a watch. Just read that read that”. It's like, I don't do reading, and I don't do reading, I don't do form filling. But I do like doing the practical parts” (Male, T12 Incomplete) | Agreement |
| Burden | *How much effort did it take to engage in REACH-SCI?*<br>6% no effort at all<br>41% a little effort<br>12% no opinion<br>35% a lot of effort<br>6% huge effort | “Not a great amount [of effort], really. It's, it's more just restructuring my hours and getting on with my life. You know” (Male, T4 Complete)<br>“Some days a lot [of effort]. Other days, not so much and it was easier. Yeah, and it got easier as the weeks went on. I found it easier to get into the routine of get up and do it set for the days” (Female, L1 Incomplete)<br>“I felt again, I just me personally, that I was letting you and the programme down and felt a bit guilty that I couldn't or wasn't giving the program, my full dedication. Because I just just was, yeah, too busy” (Female, T10 Complete) | Agreement |
|  |  | “I actually had to buy a different band because the band that was with it, I was allergic to” (Female, L4 Incomplete) |  |
| Perceived effectiveness | *REACH-SCI is effective for reducing and breaking up inactivity* | “So, you know, so I think it's, but it's yeah, but I'm definitely, I'm definitely more active now than I was. And I've definitely got more energy now than I had when I came out when I first came onto the programme” (Female, T3 Incomplete)<br>“It's come at the right time, because if, if you don't start now, you're just going to get lazier and lazier... remember, most of the time you're homebound, so it's good to be doing it now, so that you're not lazy... I want to be active now, rather than later” (Male, T3 Complete)<br>“Yeah, and it makes you think more about the inactivity, you know, having talked about, I'd never have thought about that in a negative sense. You don't think about periods of inactivity, do you? Until someone actually talks to you about it” (Male, T5 Incomplete)<br>“Yes [intervention was effective]. Erm, definitely the idea of watching TV for 30 minutes, then doing something. Within like the first two weeks, I really got into it. But I got so into it, I went to the cinema in week 6, and without me even realising, I was kind of just wheeling around. And the person I was with was kind of like “what?”, and I was like, “oh I'll sit back down”. So yeah, I got so into it that” (Female, L4 Incomplete)<br>“I think I think where this programme is more useful is for somebody who is perhaps not coping with their disability, or they have a lot more problems with their disability... And maybe they do spend a lot of time just sitting and watching telly or playing games on the computer or not doing very much” (Female, T10 Complete) | Agreement |
|  | 6% strongly disagree<br>0% disagree<br>12% no opinion<br>65% agree<br>18% strongly agree |  |  |
| Ethicality | *How fair is REACH-SCI for individuals with paraplegia?* | “No [ethical concerns], I think everything was well-organised” (Female, T5 Complete) | Agreement |
|  | 0% very unfair<br>0% unfair<br>12% no opinion<br>41% fair<br>47% very fair | “No concerns” (Male, T9 Incomplete) |  |
| Intervention coherence | *It is clear to me how REACH-SCI helps to reduce inactivity*<br>0% strongly disagree<br>0% disagree<br>6% no opinion<br>76% agree<br>18% strongly agree | “Yeah [it is clear how intervention is intended to work], and it makes you think more about the inactivity, you know, I'd never have thought about that in a in a negative sense. You don't think about periods of inactivity, do you? Until someone actually talks to you about it” (Male, T5 Incomplete)<br>“Yes. I mean, the leaflet is fairly clear on, or the booklet rather, is very clear on how it's supposed to work” (Female, T3 Incomplete)<br>“Yeah, well you explained it initially. And then when we came for the initial testing upstairs. Yeah, erm, you said this is what this is for. And then we went away with tools that you gave us, and you explained these are what we can be using in different times each day. What we can use and more in different situations” (Male, T8 Incomplete) | Agreement |
| Opportunity costs | *Taking part in REACH-SCI interfered with my other priorities or commitments*<br>24% strongly disagree<br>41% disagree<br>29% no opinion<br>6% agree<br>0% strongly disagree | “‘No, I think it was fine and fair, to be fair. I feel like it was, it was a clinical study being done in the background of my day-to-day life. It wasn't. It didn't feel like I was actually doing anything over and out of the way of my normal routines” (Female, T11 Incomplete)<br>Yes [taking part interfered with other priorities or commitments]. Yeah, and I did put that it did on the survey. But just because sometimes I have to focus quite hard at work. And I mean, obviously I wasn't working as much at the moment, but still trying to do things along the lines of that and it, it did make it quite tricky (Female, T3 Incomplete) | Partial agreement |
|  |  | “I didn’t give it 100%, I didn’t. I should have but I didn’t. But I’ve got other commitments. That had to come first as that’s life. This is, like, you’ve got other participants that are gonna give you the data you need. I’m not the only one, so it’s like, well, you know” (Male, T12 Incomplete) |  |
| Self-efficacy | *How confident did you feel about engaging with the REACH-SCI programme?*<br>0% very unconfident<br>0% not confident<br>12% no opinion<br>53% confident<br>35% very confident | “Yeah, yeah, it's pretty straightforward, and it gives you the confidence you need, and yeah” (Male, T3 Complete)<br>Mm reasonably confident reasonably. I was a bit worried I wouldn't stick with it, but I think having that those one-to-ones during the programme gave me that accountability to say “I must do it because I'm going to talk to ”, and I can't turn round and say I haven't done anything (Female, L1 Incomplete)<br>“I wasn't sure at first, but in the end, I enjoyed it. So if you need me again, give me a shout... First week was difficult. Yeah, and then you just kind of learn what you can do, and as the weeks went, it was easier. Yeah, yeah, the first week was harder” (Female, T5 Complete) | Agreement |

L, lumbar; TFA, Theoretical Framework of Acceptability; T, thoracic.

##### TFA Domain 2: Burden

Almost half of participants (47%) reported that participation required little to no effort. This was supported by qualitative findings, which indicated that participants perceived low effort was required for participation, and that participation became easier over time (Table 4; Supplementary Table 1). However, 41% of participants felt participation took a lot or a huge amount of effort, which may be explained by participants being too busy to engage. Another qualitative finding was that the wearable activity tracker band caused skin irritation in some participants, which added to the perceived burden.

##### TFA Domain 3: Perceived Effectiveness

Questionnaire data showed high perceived effectiveness, with 83% of participants agreeing or strongly agreeing that the intervention was effective. This was supported by qualitative findings, with some participants reporting that their activity levels increased and REACH-SCI was useful for newly injured individuals (Table 4; Supplementary Table 1). High perceived effectiveness is explained by participant quotes stating that the intervention increased their knowledge of sedentary behaviour and helped with the formation of new habits. However, some participants felt that the intervention is less useful for individuals who are already active.

##### TFA Domain 4: Ethicality

Quantitative data found that 88% of participants felt REACH-SCI was fair for individuals with paraplegia (Table 4; Supplementary Table 1). As such, qualitative analysis also identified that there were no ethical concerns.

##### TFA Domain 5: Intervention Coherence

Nearly all participants (94%) agreed or strongly agreed that it was clear how the intervention was intended to reduce inactivity (Table 4; Supplementary Table 1). Consistent with this, qualitative findings indicated that the educational booklet improved participants’ knowledge of sedentary behaviour, while clear written materials and researcher explanations enhanced the understanding of the intervention.

##### TFA Domain 6: Opportunity Costs

Most participants (65%) disagreed or strongly disagreed that participation in REACH-SCI interfered with other priorities or commitments (Table 4; Supplementary Table 1). This was reflected in the qualitative data with many participants reporting little interference from the intervention. However, some did feel aspects of life interfered with the intervention, such as difficulty implementing activity breaks in workplace settings. These findings suggest disagreement in the qualitative data and that differences in opportunity costs are situated according to individual circumstances.

##### TFA Domain 7: Self-efficacy

Self-efficacy was high, with 88% of participants agreeing or strongly agreeing that they felt confident in participating in REACH-SCI (Table 4). This was consistent with qualitative findings with participants describing confidence in their ability to participate in the intervention. High self-efficacy may be explained by motivational support sessions increasing confidence in participation. However, some participants noted lower self-efficacy initially but described that their confidence increased over time.

#### Acceptability/Perceived Effectiveness of Individual Intervention Components

##### Wearable Activity Tracker

Participants rated the wearable activity tracker as highly useful for breaking up sedentary behaviour (4.0 ± 1.0 out of 5; Table 5). Qualitative findings help explain this positive scoring, with participants describing the inactivity reminders as useful and tracking activity levels was motivational. However, despite opinions of this component being generally favourable, some participants reported that the inactivity reminders and daily push count data were inaccurate.

**Table 5.**
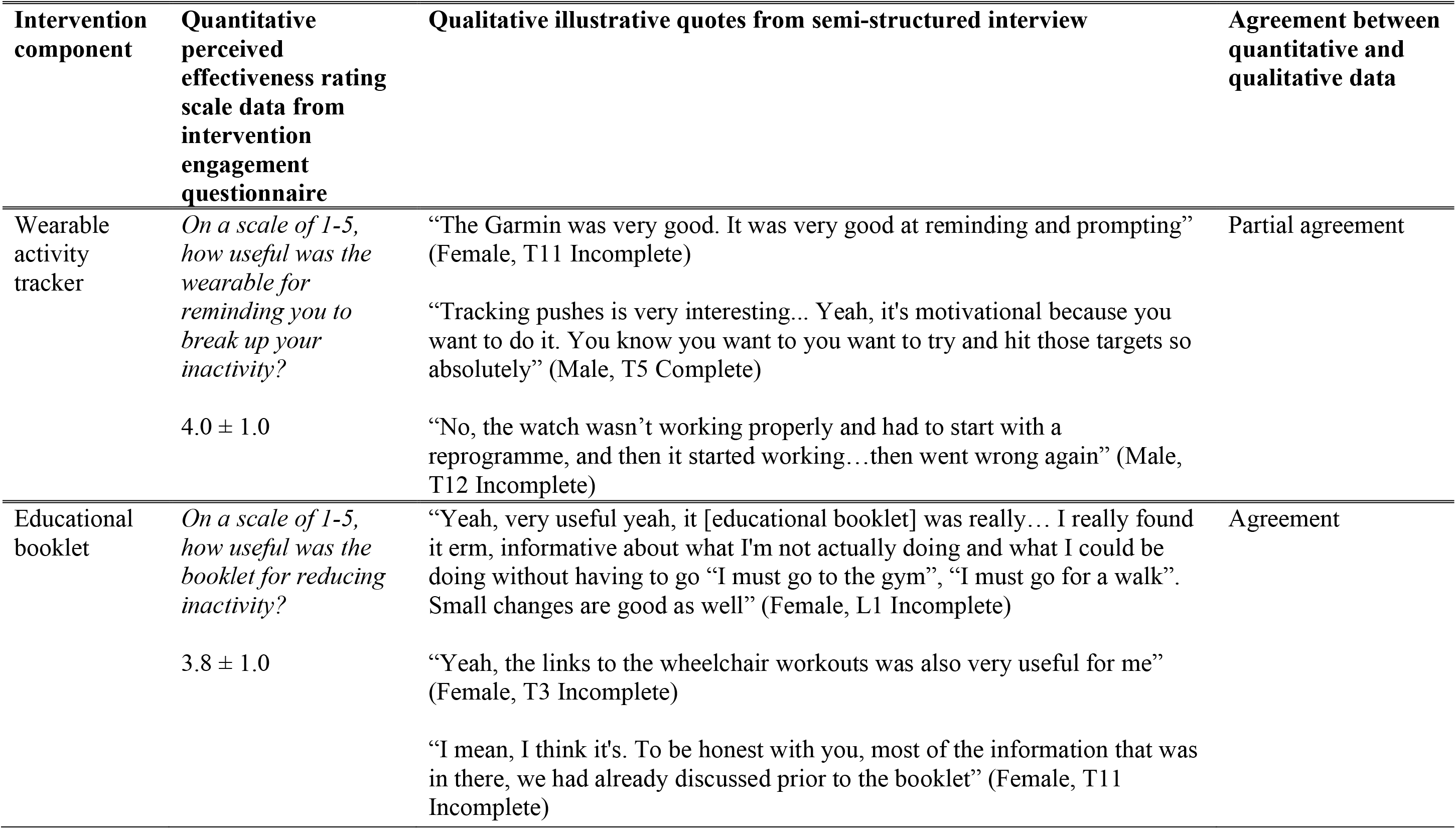
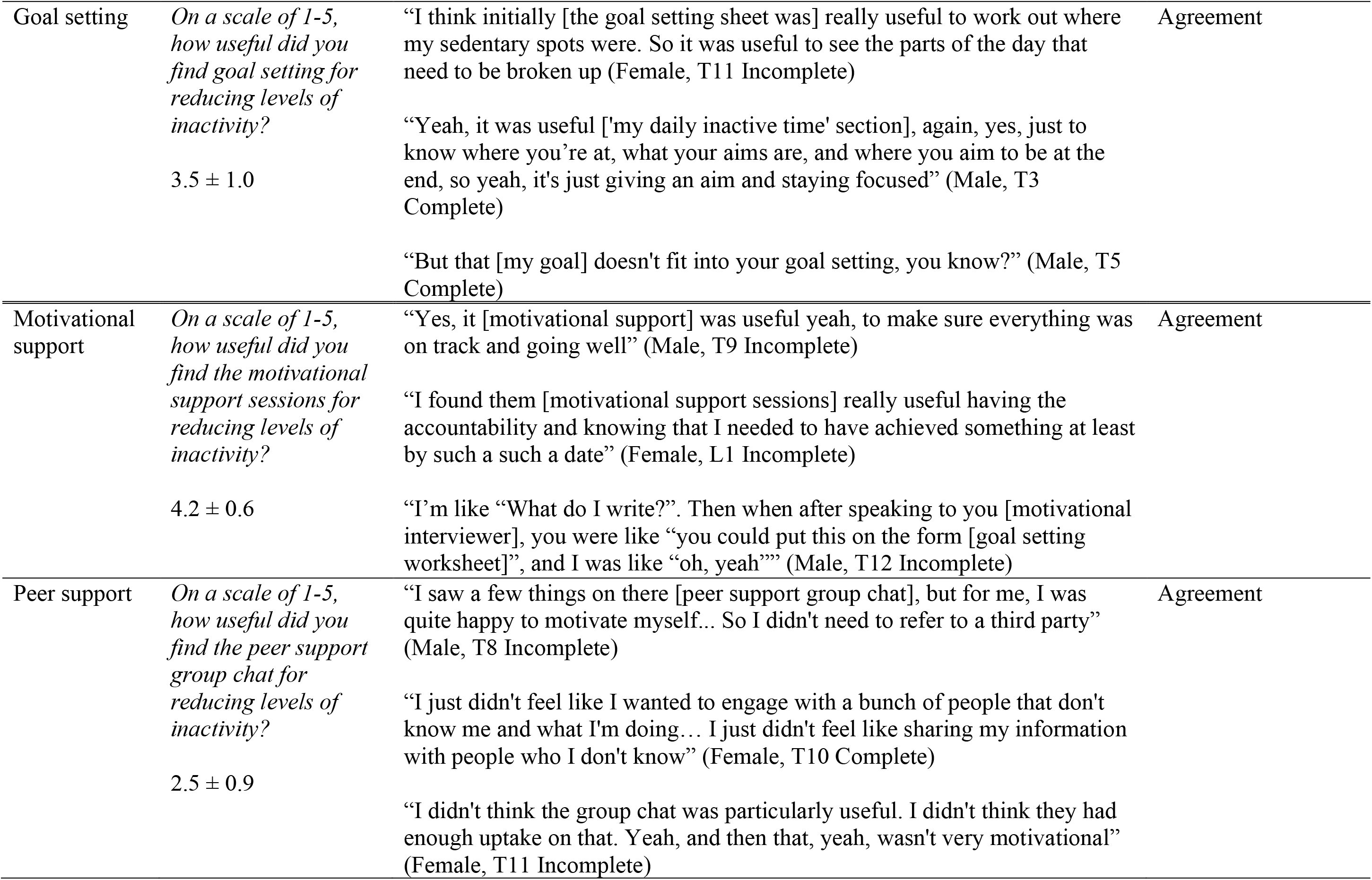
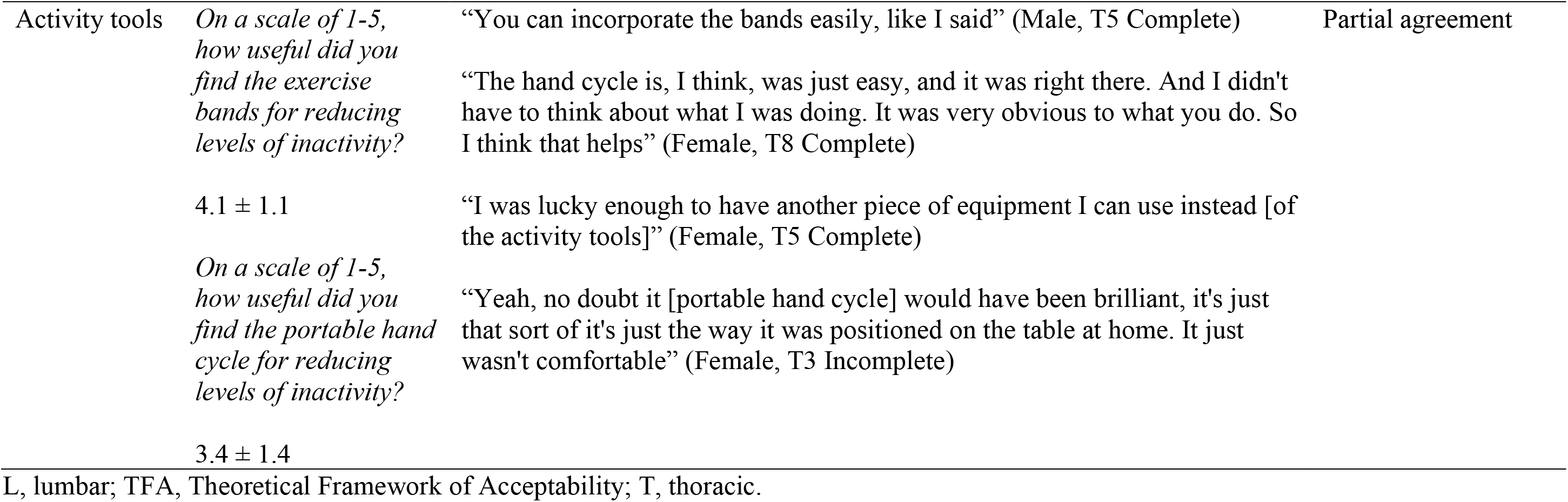
Integrated mixed-methods acceptability/perceived effectiveness findings for individual intervention components.

##### Educational Booklet

The educational booklet was rated 3.8 ± 1.0 for usefulness in reducing sedentary behaviour (Table 5). Qualitative findings help explain this positive rating, with participants describing the booklet as a useful source of information, and valued the signposting to additional resources. However, some participants reported already knowing the information.

##### Goal Setting

Goal setting was rated 3.5 ± 1.0 for its usefulness (Table 5). Qualitative findings provided insights into this rating, with participants reporting that goal setting improved their awareness of inactivity and that goal setting kept them focused and on track with the intervention. Despite the positive nature of these findings, some participants expressed that the rigid, structured format of the worksheet prevented the personalisation of goals.

##### Motivational Support

Motivational support was perceived as highly useful for reducing sedentary behaviour (4.2 ± 0.6; Table 5). Qualitative findings helped explain this positive rating with participants reporting that sessions kept them focused, accountable and on track with the intervention. Participants also valued the information and practical ideas provided by the motivational interviewer.

##### Peer Support

The peer support group received the lowest rating for usefulness of all the intervention components (2.5 ± 2.9; Table 5). Qualitative findings provided some insight into this low rating, with some participants stating that they did not need peer support, they felt uncomfortable sharing personal experiences with strangers and that there was not enough engagement from other participants in the group chat.

##### Activity Tools

Participants rated the exercise bands as useful and the portable hand cycle slightly less useful for reducing sedentary behaviour (4.1 ± 1.1 and 3.4 ± 1.4, respectively; Table 5). Qualitative data help explain these scores, with participants reporting that both tools were easy to use and incorporate into everyday routines. However, perceived effectiveness was lower for some participants who already owned similar activity tools. The lower rating for the portable hand cycle can be explained by some participants not having suitable space to use the equipment.

### Intervention Safety

Seven participants each reported one adverse event. Five participants (29%) experienced skin irritation from the wearable activity tracker, and two (12%) experienced shoulder pain. All adverse events were mild in nature. Participants with skin irritation were advised to swap the wearable activity tracker to the opposite wrist until irritation cleared, and to regularly remove and clean the wrist strap. Participants with shoulder pain were advised to stop using the activity tools if new pain occurred or existing pain worsened, and to follow the advice provided in the guidance sheet around maintaining proper form and monitoring pain, fatigue and spasticity. All symptoms resolved with the management strategies provided and participants were able to continue with the intervention as planned. No serious adverse events were reported. There was a small mean increase in generalised pain as measured by the MPQ (Δ 3.4 ± 17.9; out of 78). In contrast, there was a decrease in fatigue (Δ -9.5 ± 15.4; out of 84) as measured by the MFIS.

### Intervention Feasibility

All 17 participants who completed baseline measurements received a wearable activity tracker (or given guidance on how to use relevant features on their own device; n = 3, 18%), an educational booklet, a goal setting worksheet and exercise bands. One participant (6%) declined the portable hand cycle due to already having a similar device. Attendance at the motivational support sessions was high with 12 participants (71%) attending all three sessions, four (24%) attended two sessions, and one participant (6%) attended one session.

### Intervention Engagement

All 17 participants reported using the wearable activity tracker during the intervention. Sixteen (94%) and 15 (88%) participants used the tracker 6-7 days per week in the first and second months, respectively (Table 6). Most participants reported reading the whole educational booklet (n = 15, 88%). Engagement with each of the booklet sections was high, but less than half (44%) of participants used the links to relevant resources. Limited engagement with the links to relevant resources may have been due to the burden of reading the final section of the booklet, with one participant noting “*[It takes too much] Time and I don’t like reading*” (Male, T12 Incomplete lesion). Despite this, participants who used the links found them useful: “*Wheelchair workouts were especially useful*” (Female, T3 Incomplete lesion). All participants used the goal setting worksheet to set goals and most participants (71%) referred back to the worksheet during REACH-SCI to review their progress. Reasons for not referring back to the worksheet included “*It is repetitive and does not need repeating*” (Male, T5 Complete lesion).

**Table 6.**
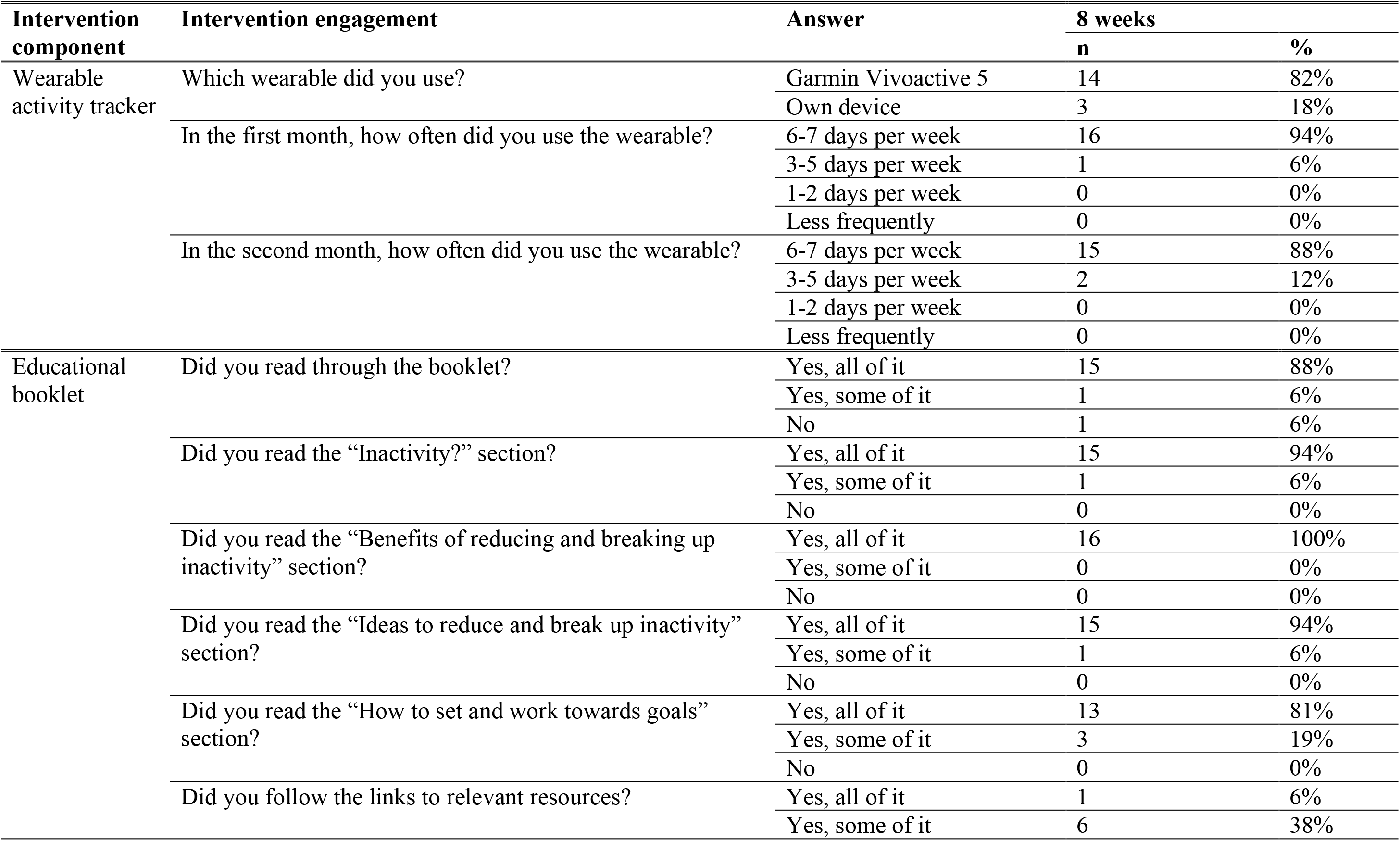
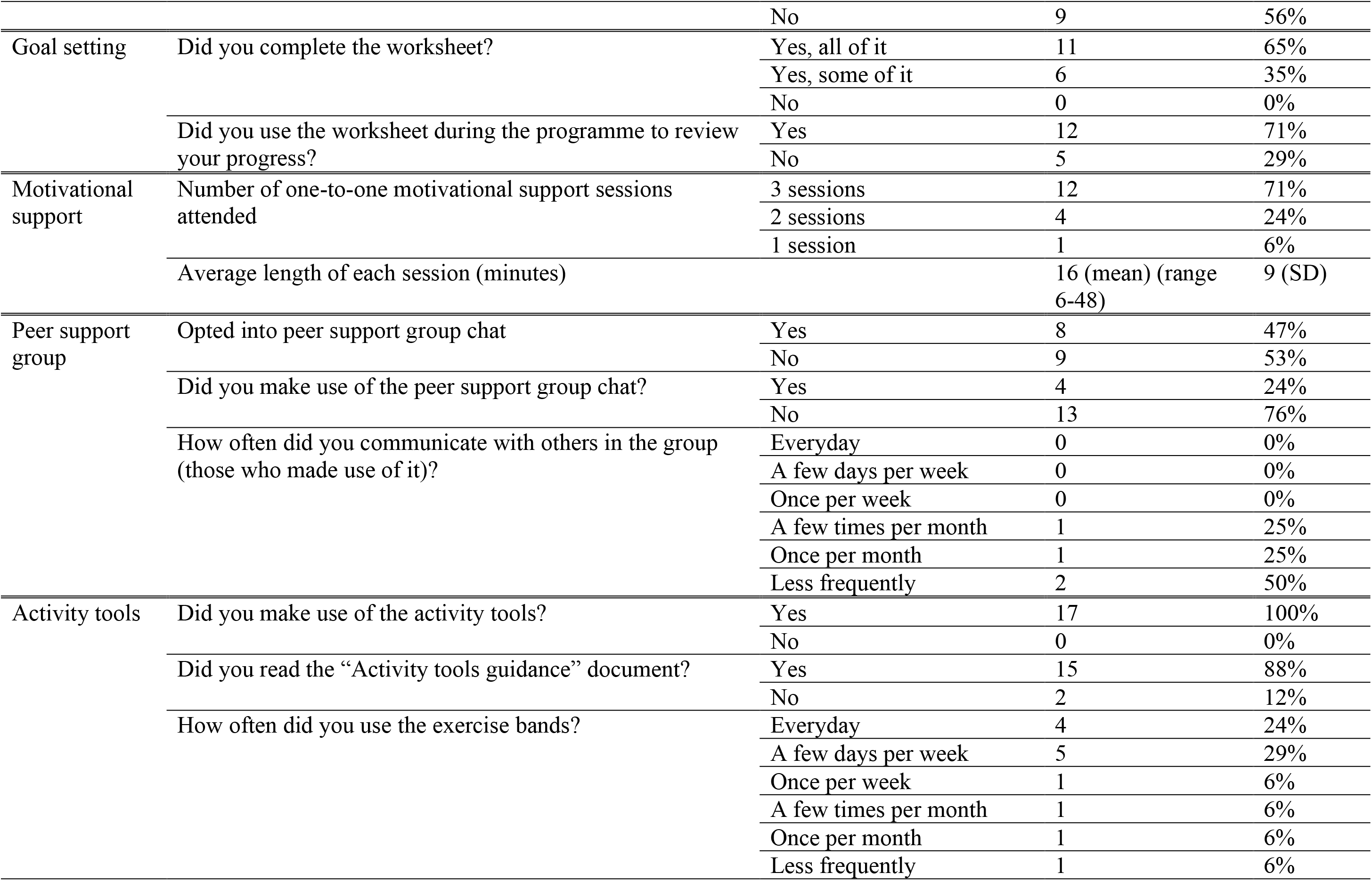
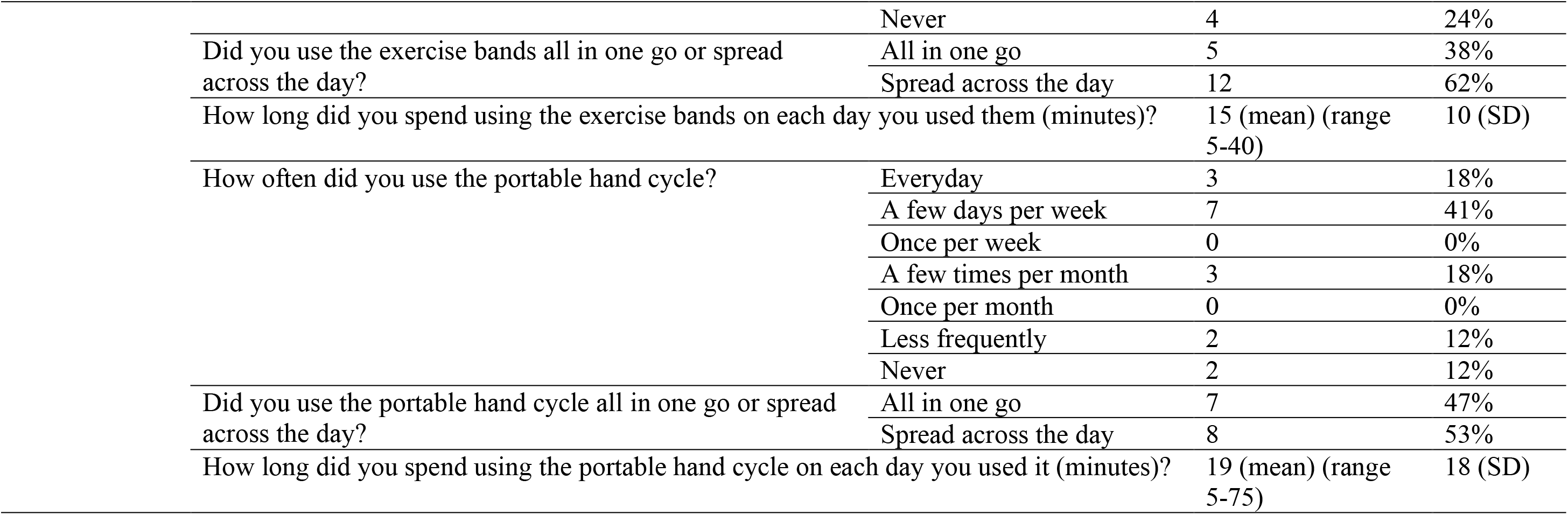
Questionnaire responses for participant engagement with intervention components.

| Intervention component | Intervention engagement | Answer | 8 weeks |  |
| --- | --- | --- | --- | --- |
|  |  |  | n | % |
| Wearable activity tracker | Which wearable did you use? | Garmin Vivoactive 5 | 14 | 82% |
|  |  | Own device | 3 | 18% |
|  | In the first month, how often did you use the wearable? | 6-7 days per week | 16 | 94% |
|  |  | 3-5 days per week | 1 | 6% |
|  |  | 1-2 days per week | 0 | 0% |
|  |  | Less frequently | 0 | 0% |
|  | In the second month, how often did you use the wearable? | 6-7 days per week | 15 | 88% |
|  |  | 3-5 days per week | 2 | 12% |
|  |  | 1-2 days per week | 0 | 0% |
|  |  | Less frequently | 0 | 0% |
| Educational booklet | Did you read through the booklet? | Yes, all of it | 15 | 88% |
|  |  | Yes, some of it | 1 | 6% |
|  |  | No | 1 | 6% |
|  | Did you read the “Inactivity?” section? | Yes, all of it | 15 | 94% |
|  |  | Yes, some of it | 1 | 6% |
|  |  | No | 0 | 0% |
|  | Did you read the “Benefits of reducing and breaking up inactivity” section? | Yes, all of it | 16 | 100% |
|  |  | Yes, some of it | 0 | 0% |
|  |  | No | 0 | 0% |
|  | Did you read the “Ideas to reduce and break up inactivity” section? | Yes, all of it | 15 | 94% |
|  |  | Yes, some of it | 1 | 6% |
|  |  | No | 0 | 0% |
|  | Did you read the “How to set and work towards goals” section? | Yes, all of it | 13 | 81% |
|  |  | Yes, some of it | 3 | 19% |
|  |  | No | 0 | 0% |
|  | Did you follow the links to relevant resources? | Yes, all of it | 1 | 6% |
|  |  | Yes, some of it | 6 | 38% |
|  |  | No | 9 | 56% |
| Goal setting | Did you complete the worksheet? | Yes, all of it | 11 | 65% |
|  |  | Yes, some of it | 6 | 35% |
|  |  | No | 0 | 0% |
|  | Did you use the worksheet during the programme to review your progress? | Yes | 12 | 71% |
|  |  | No | 5 | 29% |
| Motivational support | Number of one-to-one motivational support sessions attended | 3 sessions | 12 | 71% |
|  |  | 2 sessions | 4 | 24% |
|  |  | 1 session | 1 | 6% |
|  | Average length of each session (minutes) |  | 16 (mean) (range 6-48) | 9 (SD) |
| Peer support group | Opted into peer support group chat | Yes | 8 | 47% |
|  |  | No | 9 | 53% |
|  | Did you make use of the peer support group chat? | Yes | 4 | 24% |
|  |  | No | 13 | 76% |
|  | How often did you communicate with others in the group (those who made use of it)? | Everyday | 0 | 0% |
|  |  | A few days per week | 0 | 0% |
|  |  | Once per week | 0 | 0% |
|  |  | A few times per month | 1 | 25% |
|  |  | Once per month | 1 | 25% |
|  |  | Less frequently | 2 | 50% |
| Activity tools | Did you make use of the activity tools? | Yes | 17 | 100% |
|  |  | No | 0 | 0% |
|  | Did you read the “Activity tools guidance” document? | Yes | 15 | 88% |
|  |  | No | 2 | 12% |
|  | How often did you use the exercise bands? | Everyday | 4 | 24% |
|  |  | A few days per week | 5 | 29% |
|  |  | Once per week | 1 | 6% |
|  |  | A few times per month | 1 | 6% |
|  |  | Once per month | 1 | 6% |
|  |  | Less frequently | 1 | 6% |

|  |  |  |  |
| --- | --- | --- | --- |
|  | Never | 4 | 24% |
| Did you use the exercise bands all in one go or spread across the day? | All in one go | 5 | 38% |
|  | Spread across the day | 12 | 62% |
| How long did you spend using the exercise bands on each day you used them (minutes)? |  | 15 (mean) (range 5-40) | 10 (SD) |
| How often did you use the portable hand cycle? | Everyday | 3 | 18% |
|  | A few days per week | 7 | 41% |
|  | Once per week | 0 | 0% |
|  | A few times per month | 3 | 18% |
|  | Once per month | 0 | 0% |
|  | Less frequently | 2 | 12% |
|  | Never | 2 | 12% |
| Did you use the portable hand cycle all in one go or spread across the day? | All in one go | 7 | 47% |
|  | Spread across the day | 8 | 53% |
| How long did you spend using the portable hand cycle on each day you used it (minutes)? |  | 19 (mean) (range 5-75) | 18 (SD) |

Engagement with the motivational support sessions was generally high. A reason for non-engagement was “*I felt able to self-motivate*” (L1 Incomplete, Male). Less than half of participants (47%) opted into the peer support group chat, with reasons for non-use of this component including “*I prefer to work on my own and wasn’t interested in sharing my regime with other people*” (T10 Complete, Female). Low levels of engagement amongst those that did opt into the group chat included “*Not much input from other users*” (T5 Complete, Male). All participants used at least one of the activity tools. Over half of participants used the exercise bands (53%) and portable hand cycle (59%) at least a few times per week.

### Acceptability of Data Collection Procedures

Participants generally had a positive view of the data collection procedures. The burden of data collection was deemed low: “*Minimal really, there’s not really a lot of effort involved [in the measurements]*” (Male, T9 Complete). Additionally, the flexibility of data collection sessions around participants’ routines was noted: “*You know, you work with me, on trying to fit it into the best time of day*” (Male, T5 Incomplete). Another positive finding identified was the accessibility of the facilities where the measurements took place: “*You’ve got a good set up… This height adjustable bed… it was really helpful*” (Female, T5 Complete). Some participants found that taking part in the measurements was interesting: “*I find it fascinating coming here and I like being around the academia*” (Male, L1 Incomplete).

Participants identified several factors that could affect the acceptability of study procedures. Travel requirements were seen as burdensome, with one reporting that: “*Only coming here and coming back [was an effort]*” (Male, T8 Incomplete). Concerns were also raised around the safety of one data collection procedure: “*Getting on to those scales can be very yeah, a bit risky*” (Female, L4 Incomplete). In addition, some participants highlighted concerns around the potentially sensitive nature of the mental health questionnaires: “*The PHQ and GAD I can imagine if someone were struggling with their mood, that might be tricky*” (Female, T8 Complete).

### Study Feasibility

Sixty potential participants were identified through contacting previous study participants (n = 42), social media posts (n = 9), the SCI trials finder website (n = 5), the partner NHS site (n = 3) and snowballing (n = 1). A total of 30 individuals expressed an interest and were screened for eligibility (Figure 2). Twelve participants were excluded due to not meeting the inclusion criteria. All 18 individuals deemed eligible (60% eligibility rate) were enrolled into the study (100% recruitment rate). Seventeen participants received the intervention and completed the study (94% retention rate), whilst one was withdrawn before baseline measurements due to loss of contact (6% attrition rate). All 17 participants who completed the study had a complete, valid dataset for each outcome measure (100% data completion rate).

### Preliminary Efficacy

#### Sedentary Behaviour and Physical Activity

All sedentary behaviour outcomes showed descriptive improvements in mean values (Table 7). Descriptive increases were also observed in light intensity physical activity, MVPA and total physical activity.

**Table 7.** Preliminary efficacy of the intervention. Data presented as mean ± SD.

| **Outcome** | **Baseline** | **8 weeks** | **Mean change** |
| --- | --- | --- | --- |
| **Sedentary behaviour and physical activity** |  |  |  |
| Sedentary time (min/day) | 748.2 ± 175.4 | 717.9 ± 153.3 | -30.3 ± 83.2 |
| Breaks in sedentary behaviour (number/day) | 36.1 ± 20.0 | 39.7 ± 27.7 | 3.6 ± 16.9 |
| Percent of day in sedentary behaviour | 77.0 ± 11.4 | 75.2 ± 12.1 | -1.8 ± 7.4 |
| Time in prolonged sedentary bouts > 30 min (min/day) | 561.7 ± 254.9 | 519.0 ± 263.1 | -42.7 ± 133.2 |
| Time in prolonged sedentary bouts > 60 min (min/day) | 469.0 ± 267.9 | 432.8 ± 278.8 | -36.3 ± 155.8 |
| Number of prolonged sedentary bouts > 30 min | 7.0 ± 2.5 | 5.8 ± 1.9 | -1.2 ± 2.7 |
| Number of prolonged sedentary bouts > 60 min | 4.1 ± 1.5 | 3.4 ± 1.4 | -0.7 ± 1.5 |
| Light intensity physical activity (min/day) | 205.4 ± 86.4 | 219.9 ± 105.7 | 14.5 ± 65.4 |
| MVPA (min/day) | 9.7 ± 7.5 | 15.2 ± 15.1 | 5.5 ± 10.9 |
| Total physical activity (min/day) | 215.2 ± 91.2 | 235.2 ± 111.9 | 20.0 ± 69.2 |
| **Cardiovascular disease biomarkers** |  |  |  |
| Systolic blood pressure (mmHg) | 122.9 ± 14.7 | 121.2 ± 15.4 | -1.6 ± 12.9 |
| Diastolic blood pressure (mmHg) | 79.5 ± 12.2 | 77.2 ± 13.5 | -2.3 ± 9.2 |
| Mean arterial pressure (mmHg) | 93.8 ± 12.2 | 91.9 ± 13.3 | -1.9 ± 9.4 |
| Body mass (kg) | 83.7 ± 18.6 | 83.2 ± 18.8 | -0.5 ± 2.2 |
| Body mass index (kg/m<sup>2</sup>) | 28.3 ± 7.2 | 28.1 ± 7.0 | -0.2 ± 0.8 |
| Waist circumference (cm) | 97.7 ± 14.1 | 96.7 ± 14.5 | -1.0 ± 3.2 |
| Body fat mass (kg) | 29.4 ± 13.0 | 30.8 ± 13.1 | 1.4 ± 3.6 |
| Body fat percentage (%) | 34.2 ± 9.1 | 36.4 ± 8.8 | 2.2 ± 4.7 |
| Total cholesterol (mmol/L) | 5.2 ± 1.4 | 4.9 ± 1.2 | -0.2 ± 0.74 |
| HDL cholesterol (mmol/L) | 1.4 ± 0.4 | 1.4 ± 0.4 | -0.0 ± 0.2 |
| Non-HDL cholesterol (mmol/L) | 3.8 ± 1.3 | 3.5 ± 1.1 | -0.2 ± 0.7 |
| Total cholesterol:HDL cholesterol ratio | 3.9 ± 1.4 | 3.7 ± 1.3 | -0.1 ± 0.6 |
| Glucose (mmol/L) | 5.6 ± 0.7 | 5.7 ± 0.9 | 0.1 ± 0.6 |
| **Psychosocial outcomes** |  |  |  |
| Anxiety | 4.6 ± 4.6 | 3.2 ± 2.9 | -1.5 ± 5.0 |
| Depression | 7.5 ± 5.1 | 5.2 ± 4.1 | -2.4 ± 6.0 |
| Wellbeing: Life satisfaction | 5.7 ± 1.9 | 6.3 ± 1.5 | 0.6 ± 2.3 |
| Wellbeing: Life worthwhile | 6.3 ± 2.0 | 6.8 ± 2.2 | 0.5 ± 3.1 |
| Wellbeing: Happiness | 6.7 ± 2.0 | 6.5 ± 2.1 | -0.2 ± 2.7 |
| Wellbeing: Anxiety | 3.6 ± 2.4 | 2.6 ± 2.3 | -0.9 ± 2.9 |
| Health-related quality of life: Physical functioning | 39.7 ± 22.8 | 42.4 ± 17.5 | 3.2 ± 25.5 |
| Health-related quality of life: Physical role functioning | 44.1 ± 39.1 | 58.8 ± 37.4 | 14.7 ± 45.1 |
| Health-related quality of life: Emotional role functioning | 66.7 ± 42.5 | 74.5 ± 34.4 | 7.8 ± 30.1 |
| Health-related quality of life: Vitality | 46.5 ± 14.6 | 54.7 ± 15.3 | 8.2 ± 19.4 |
| Health-related quality of life: Mental health | 69.6 ± 19.5 | 74.8 ± 18.5 | 5.2 ± 23.4 |
| Health-related quality of life: Social functioning | 64.7 ± 32.2 | 77.2 ± 20.8 | 12.5 ± 32.2 |
| Health-related quality of life: Bodily pain | 49.1 ± 25.9 | 67.6 ± 32.8 | 18.5 ± 36.0 |
| Health-related quality of life: General health | 46.2 ± 25.2 | 53.2 ± 23.8 | 7.1 ± 28.4 |
| Health-related quality of life: Physical component summary | 32.6 ± 9.4 | 37.0 ± 9.0 | 4.5 ± 11.9 |
| Health-related quality of life: Mental component summary | 50.0 ± 11.8 | 53.1 ± 9.2 | 3.1 ± 10.0 |
| Fatigue | 36.7 ± 16.6 | 27.2 ± 14.4 | -9.5 ± 15.4 |
| Pain | 38.7 ± 16.3 | 42.1 ± 21.1 | 3.4 ± 17.9 |

HDL, high-density lipoprotein; MVPA, moderate-to-vigorous intensity physical activity.

#### Cardiovascular Disease Biomarkers

Descriptive analysis demonstrated mean reductions in systolic blood pressure, diastolic blood pressure, MAP, body mass, BMI, waist circumference, total cholesterol, non-HDL cholesterol and total cholesterol:HDL cholesterol ratio (Table 7). HDL cholesterol remained unchanged, while body fat mass and body fat percentage showed descriptive increases.

#### Psychosocial Outcomes

Descriptive improvements were observed in mean anxiety and depression scores, some wellbeing domains (life satisfaction, life being worthwhile and anxiety), and all health-related quality of life domains and summary scores (Table 7).

## Discussion

This study’s mixed-methods analysis demonstrated that the REACH-SCI intervention was acceptable to wheelchair-users with paraplegia across all constructs of the TFA. The high levels of acceptability of REACH-SCI may in part be due to the co-design approach, which ensured the intervention components were tailored to this population’s needs. The acceptability of REACH-SCI is consistent with the evidence-informed approach to physical activity counselling that recommends starting with lower-intensity activities of daily living to achieve long-term health behaviour change in individuals with physical impairments, such as SCI [42]. Furthermore, acceptability of REACH-SCI that combined a wearable activity tracker, education, goal setting and one-to-one motivational support sessions, is also consistent with findings from studies incorporating similar components in individuals with neurological conditions such as stroke [43] and multiple sclerosis [44]. Together, these findings suggest that sedentary behaviour is an acceptable intervention target in individuals with paraplegia and that the REACH-SCI intervention warrants further evaluation to determine its effectiveness.

Participants suggested several areas for improvements to REACH-SCI. A reduction in the amount of paperwork to be completed was a frequent suggestion, and that more flexible and accessible ways to engage, such as having electronic forms, websites, or video materials could be incorporated in future iterations of the intervention. Another recurring suggestion was that other commitments interfered with intervention engagement, which could be overcome by focusing more on lifestyle barriers to participation in motivational support sessions, such as workplace restrictions. In addition, some participants described having a lack of confidence at the start of the intervention, which could be helped by earlier provision of motivational support on day 1 of the intervention. Together, these findings provide valuable insights for future refinements of the intervention, and may enhance both the acceptability and potential effectiveness of REACH-SCI.

Consistent with previous sedentary behaviour interventions in individuals with neurological conditions [45], REACH-SCI was safe with no serious adverse events and also showed potential to reduce fatigue. Although pain increased slightly, this change was small. Shoulder pain is a common occurrence in individuals with SCI due to overuse for mobility [46] and advice around appropriate shoulder protection was provided with the activity tools. Therefore, any increase in pain may not have been attributed to the intervention. The findings observed in the current study suggest that, overall, REACH-SCI is safe for individuals with paraplegia, but will need confirming in a larger effectiveness study.

In line with a sedentary behaviour intervention involving older adults with frailty [47], feasibility of delivering components as intended and participant engagement were high for the wearable activity tracker, educational booklet, goal setting and motivational support sessions. All participants in the present study used at least one of the activity tools with over half using the exercise bands and portable hand cycle at least a few times per week. These are promising results, especially as levels of engagement were higher than those reported in a sedentary behaviour intervention in non-disabled participants who used a portable pedal machine on 37% of intervention days [48]. The high levels of participant engagement observed in REACH-SCI may reflect the co-design approach to the intervention development, meaning the components were tailored to the preferences and needs of the target population. These findings suggest that the REACH-SCI intervention components are a promising mechanism for supporting behaviour change in individuals with paraplegia.

Social facilitation was identified within a systematic review as being a key component in sedentary behaviour interventions for clinical populations [49]. However, this finding was not supported in the present study nor in an intervention with older adults [47], where uptake and engagement with peer support was low. The effectiveness of social support in Nieste et al. [49] could be due to the delivery modes employed in the included studies comprising social comparison of activity facilitated by an app/website or in-person group activities, as opposed to a group messaging app in REACH-SCI. In addition, a member of the research team had access to the group chat in REACH-SCI, which may have discouraged peer interaction [49]. In contrast, a physical activity intervention in cancer survivors involving dyadic participant-participant pairings to provide peer support had higher levels of engagement [50]. Therefore, peer support delivered through group messaging apps could be less likely to work in sedentary behaviour interventions, meaning other approaches should be explored.

This study had high eligibility, recruitment, retention and data completion rates, indicating that it is feasible to deliver and evaluate the REACH-SCI intervention. High feasibility is consistent with previous studies of sedentary behaviour interventions in individuals with long-term conditions such as multiple sclerosis [44], stroke survivors [43] and Type 2 diabetes [51]. However, the current study lacked a control arm, meaning the feasibility and acceptability of randomisation to groups is unclear. That said, similar studies have found randomised controlled designs to be feasible and acceptable [45,51].

Potential efficacy of REACH-SCI was supported through descriptive reductions in sedentary time (-30 min/day). Larger descriptive reductions in sedentary time have been reported following a four-month intervention that aimed to decrease sedentary time through the accumulation of activity across the day in individuals with paraplegia (118 min/day) [52]. However, direct comparison between studies is difficult as Martinez et al. [52] used heart rate zones to estimate sedentary behaviour, which is not a validated technique for this outcome, therefore limiting confidence in this finding. Furthermore, the present intervention demonstrated a descriptive increase in the number of breaks in sedentary behaviour (3.6 breaks/day), potentially due to the regular inactivity reminders from the wearable activity tracker. Interruptions in sedentary behaviour are independently associated with improvements in CVD biomarkers and further strengthened when combined with reductions in sedentary time [53]. This descriptive increase in sedentary breaks is greater than the mean change reported in participants with other neurological conditions, such as multiple sclerosis (1.2 breaks/day), following an intervention involving a wearable activity tracker, education, goal setting and one-to-one motivational support sessions [44]. Although comparisons across studies have to be interpreted with caution, findings suggest that REACH-SCI has potential for reducing and breaking up sedentary behaviour, supporting further evaluation in a future effectiveness trial.

Although physical activity was not specifically targeted by REACH-SCI, there was a descriptive increase in MVPA of 38.5 minutes per week. This may reflect the signposting to online resources for physical activity and exercise opportunities within the educational booklet, and the provision of activity tools, which may have encouraged the replacement of sedentary behaviour with higher intensity activities. These findings indicate that REACH-SCI could be a promising intervention for increasing physical activity alongside reducing and breaking up sedentary behaviour, which could help optimise cardiovascular-related benefits [54].

Despite being initial findings derived from a small cohort, descriptive reductions in blood pressure and waist circumference are consistent with findings from a meta-analysis of sedentary behaviour interventions in clinical populations [49]. However, there were descriptive increases in body fat mass and body fat percentage, at odds with a potential reduction in waist circumference. It is plausible, therefore, that REACH-SCI may benefit central adiposity, which is more closely related to cardiometabolic health [55] than overall body fat measurements. As previously mentioned, these are preliminary findings in a small cohort but do support the need for a definitive trial to evaluate effects on CVD biomarkers.

There was little descriptive change in fasting lipids and glucose concentrations, which is consistent with findings from a meta-analysis of sedentary behaviour interventions in clinical populations [49]. This may, in part, be explained by the generally healthy baseline lipid profiles of the participants in the present study [56]. Furthermore, there were significant improvements in long-term glucose control (as measured by glycated haemoglobin A1C) in the meta-analysis by Nieste et al. [49], suggesting that HbA1C should be included as a more sensitive biomarker when evaluating the effects of sedentary behaviour interventions.

Several psychosocial outcomes showed descriptive improvements in the current study including anxiety and depression, as well as domains of wellbeing and health-related quality of life. These preliminary findings are consistent with previous sedentary behaviour interventions in individuals with neurological conditions [43,57], potentially showing support for reduced sedentary behaviour having potential benefits beyond physical health. The largest mean change was observed in the bodily pain health-related quality of life domain, which may be important in individuals with paraplegia who experience higher levels of pain than the general population [58]. Together, these initial findings suggest that REACH-SCI could potentially benefit psychosocial health and pain, therefore warranting further effectiveness evaluation.

This study followed published guidelines for conducting and reporting feasibility trials and mixed-methods studies [18,19], as well as ensuring prospective registration of the protocol to enhance rigour, transparency and reproducibility. Furthermore, the integration of the mixed-methods data provided a greater understanding of intervention acceptability, with qualitative findings helping to contextualise and explain quantitative results. Limitations include the sample mainly consisting of White British participants, which may reduce the transferability to individuals from other ethnic backgrounds. Furthermore, the sample included a small proportion of individuals within the first year of initial discharge from inpatient rehabilitation (n = 2, 11%), limiting insight obtained from individuals at this stage of SCI rehabilitation. It is recommended that future research includes a more diverse sample so that transferability is improved.

## Conclusion

The findings of this study provide novel insights into the development and delivery of sedentary behaviour interventions for individuals with paraplegia. REACH-SCI demonstrated high levels of acceptability, safety and feasibility, with strong levels of engagement across most intervention components including the wearable activity tracker, educational booklet, goal setting, motivational support and activity tools. However, the peer support component was perceived as less acceptable, suggesting that alternative approaches to delivery modes should be considered in future iterations. The study also provided evidence of potential benefits of the intervention on behavioural, physiological and psychosocial outcomes. Overall, the findings support the progression to a definitive randomised controlled trial to determine the clinical effectiveness of REACH-SCI.

## Data Availability

Additional qualitative acceptability data can be found in Supplementary Table 1.

## **Supplementary File 1.** REACH-SCI Semi-structured Interview Guide

### Introduction

The aim of this interview is to discuss your experiences of the REACH-SCI programme, and any feedback you may have. The interview will last approximately 45 minutes. There are no right or wrong answers, it is just your opinions that count. We would like to know what worked well and what did not work well, so we can make improvements to the programme So, please feel free to speak as honestly and openly as possible. Please remember that you can stop or pause the interview at any time or chose not to answer any questions you don’t wish to. This will not affect the care you are having nor the care you receive in the future.

Just to remind you, this interview will be recorded. This is so I don’t have to write everything down, meaning I can give you my full attention. The recording will be written up word-for-word by the research team with your details anonymised. The audio file will be deleted at this point. Thanks for taking part.

### Taking part in the REACH-SCI programme

1. Did you enjoy taking part in the programme?

a. What part did you find most enjoyable, if any?
b. What part did you find least enjoyable, if any?
c. What would you suggest for making the programme more enjoyable?
2. How much effort was needed to take part in the programme?

a. Which part was the easiest or hardest to engage with and why?
b. How could we make it easier to take part in REACH-SCI?
3. Did taking part in the programme raise any ethical concerns such as privacy or safety?

a. Were any specific parts of the programme more or less concerning than others?
b. How could these be changed to avoid any concerns in the future?
4. Was the programme effective for reducing and breaking up inactivity? Why?

a. Which features of the programme were most or least effective and why?
b. How could the effectiveness of the programme be improved?
5. Was it clear to you *how* the programme was meant to help reduce and break up inactivity?

a. If it was clearer, how would it have affected your behaviour and engagement with the programme?
b. How could we make it clearer to participants?
6. How confident did you feel about taking part in the programme?

a. Were there any features of the programme you felt more or less confident about interacting with?
b. How could confidence in engaging with the programme be improved?
7. Did using the programme interfere with your other priorities or commitments?

a. E.g. length of programme, amount of content, time taken to do each one
b. Were there any parts of the programme you felt were more or less of an interference?
c. What could be changed to minimise this interference?

### Wearable activity tracker

Now I am going to ask some questions about your experience with using the wearable activity tracker.

Prompts:

1. Did you use the wearable activity tracker, and if not, why?
2. If so, was there anything you liked or disliked about it, and why?

a. What did you like or dislike about:

i. Reminders to break up inactivity?
ii. Tracking activity levels (wheelchair pushes, intensity minutes)?
iii. Rewards (streaks, badges, praise)?
iv. E.g. Ease of use, comfort, features
3. What about the wearable activity tracker would you change, and why?

### Educational resource

Now I am going to ask for your thoughts on the educational booklet.

Prompts:

1. Did you use the educational resource, and if not, why?
2. If so, how useful was the content?

a. Related to:

i. Inactivity
ii. Goal setting
iii. Techniques to maintain good health and well-being
iv. Links and contact details of relevant charities, organisations and websites
v. E.g. Amount of content, easy to understand, easy to navigate, use of written text, graphics, worksheets
3. What would you change about it and why?

### Goal setting

Now I am going to ask you about the goal setting part of the programme.

Prompts:

1. Did you use the goal setting worksheet, and if not, why?
2. If so, how useful was the goal setting worksheet?

a. Related to:

i. The “my inactivity” section
ii. The “my goal setting” section
iii. The “things to consider” section
iv. The “my evaluation” section
b. E.g. Content, length, format, ease of use, instructions
3. Is there anything you would change about this part of the programme?

### Motivational support sessions

Now I am going to ask about your experiences of the motivational support sessions.

Prompts:

1. Did you do all three sessions, and if not, why?
2. How did you get on with the following aspects?

a. Topics covered
b. Regularity/length of sessions
c. Usefulness of advice/support
3. How could motivational support be improved, if at all, if REACH-SCI was to happen again?

### Peer support group chat

Now I am going to ask about your experience of the peer support group chat.

Prompts:

1. Did you opt into the group chat, and if not, why?
2. If so, how did you find specific aspects?

a. Comfort with sharing experiences within the group
b. Ease of use
c. Regularity of messages/notifications
d. Usefulness of posts/content
3. How could the peer support be improved going forward?

### Activity tools

Now I am going to ask how you found using the activity tools.

Prompts:

1. Did you use the activity tools, and if not, why?
2. If so, when did you use the activity tools and why?
3. Were there any parts of the activity tools that you particularly liked or disliked?

a. Specific piece of equipment
b. Instructions
4. Would you suggest anything different about the activity tools to improve it in the future?

### Measurement sessions

At the start and the end of the study, you took part in a series of measurements, including wearing an activity tracker, laboratory measurements, questionnaires and this interview. We would like to hear about your experiences of taking part in these measurements, so we know what worked well and what did not, so we can make improvements in the future.

Now I am going to ask about your thoughts around the measurements that were taken as part of the study.

Prompts:

1. Were there any specific measurements that you have any particular thoughts on, either positively or negatively?
2. How much effort did it take to take part in the measurement sessions?

a. Did any measurements take a lot more or less effort than the others?
3. Were there any measurements taken that you felt were problematic or unethical in any way?

a. E.g. Painful or unsafe physical measurements, distressing questions or topics, privacy concerns
4. Do you understand why we are taking these measurements from you, and how they are useful?
5. Was taking part in the measurement sessions too much of a burden?

a. Did taking part in these sessions affect you financially? How?
b. What did you think about how long the measurement sessions took?
c. Were there any issues with attendance requirements (travel and/or parking)?

### Conclusion

1. Is there anything else you would like to add?

Check participant is feeling OK at the end of this and signpost them to support services if needed (Support groups or GP).

Thank the participant again.

---

**Supplementary Table 1.**
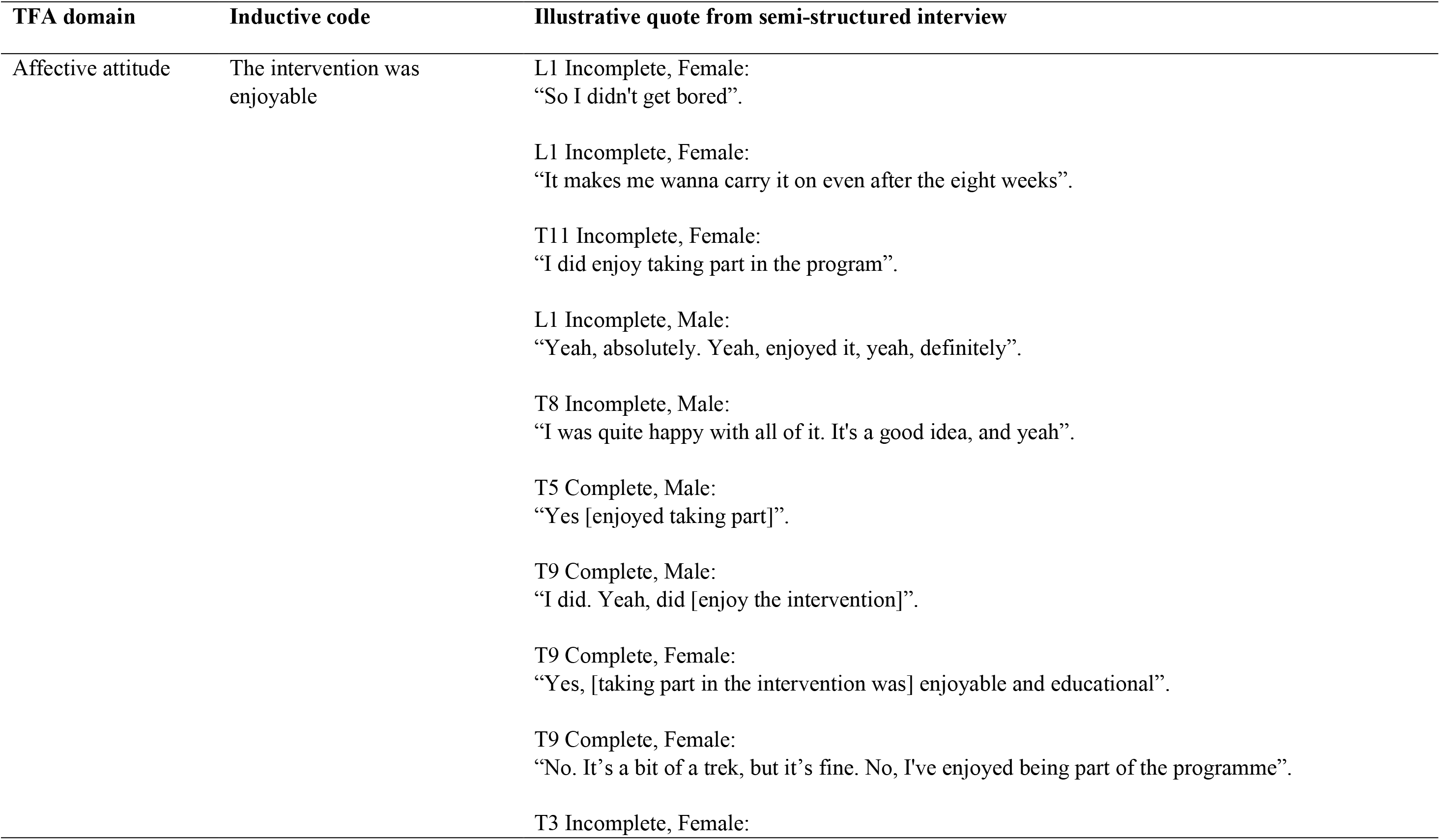
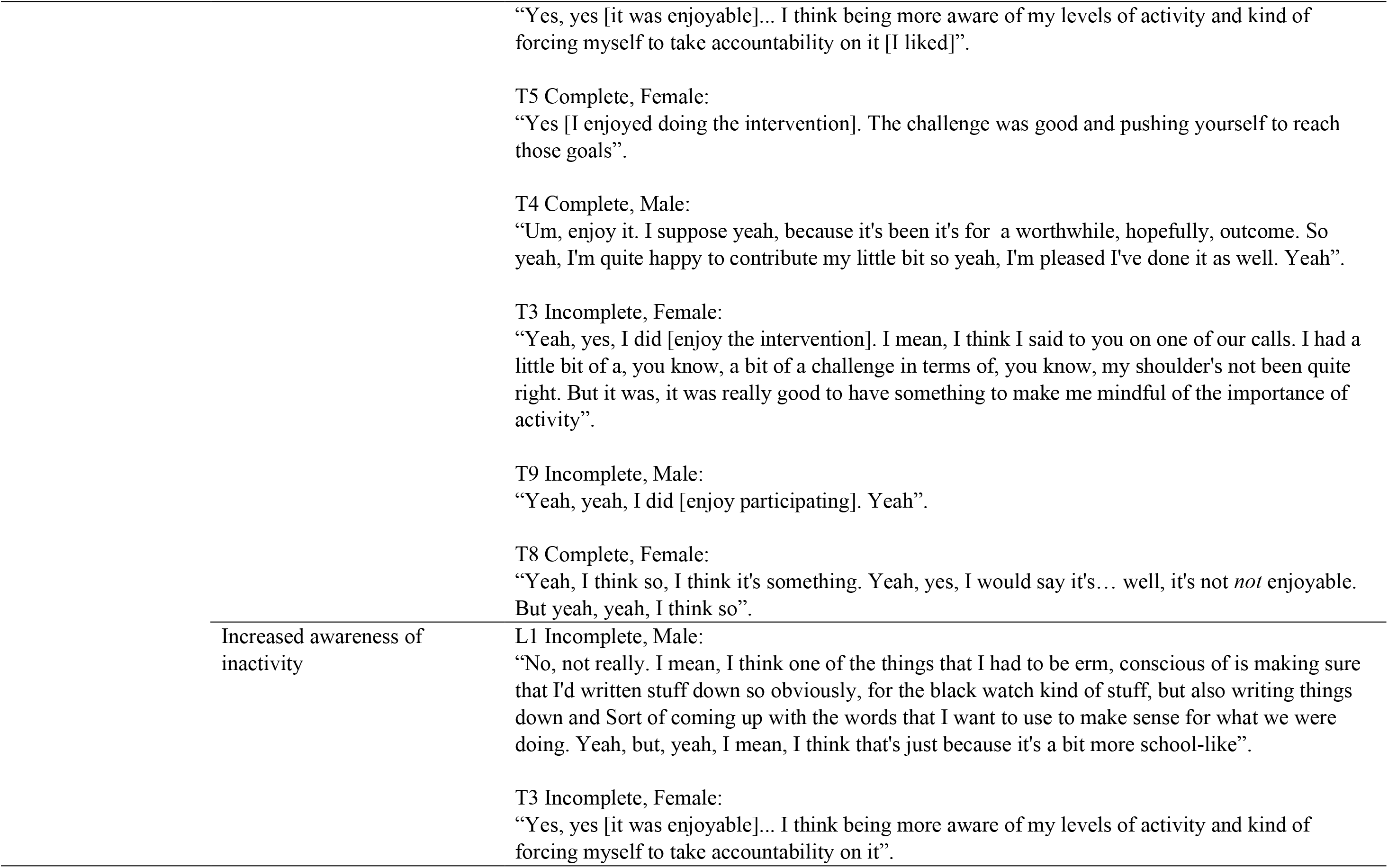
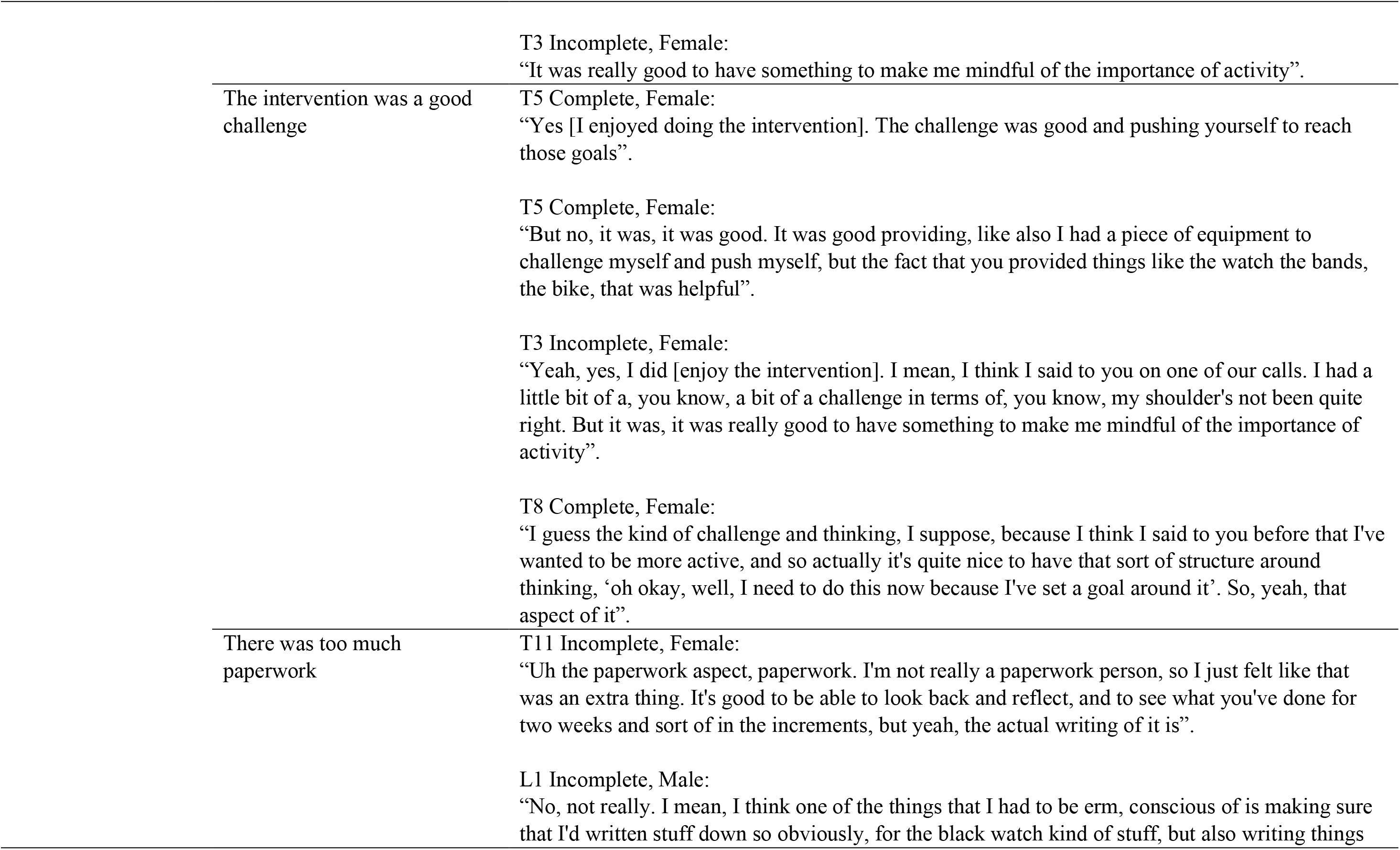
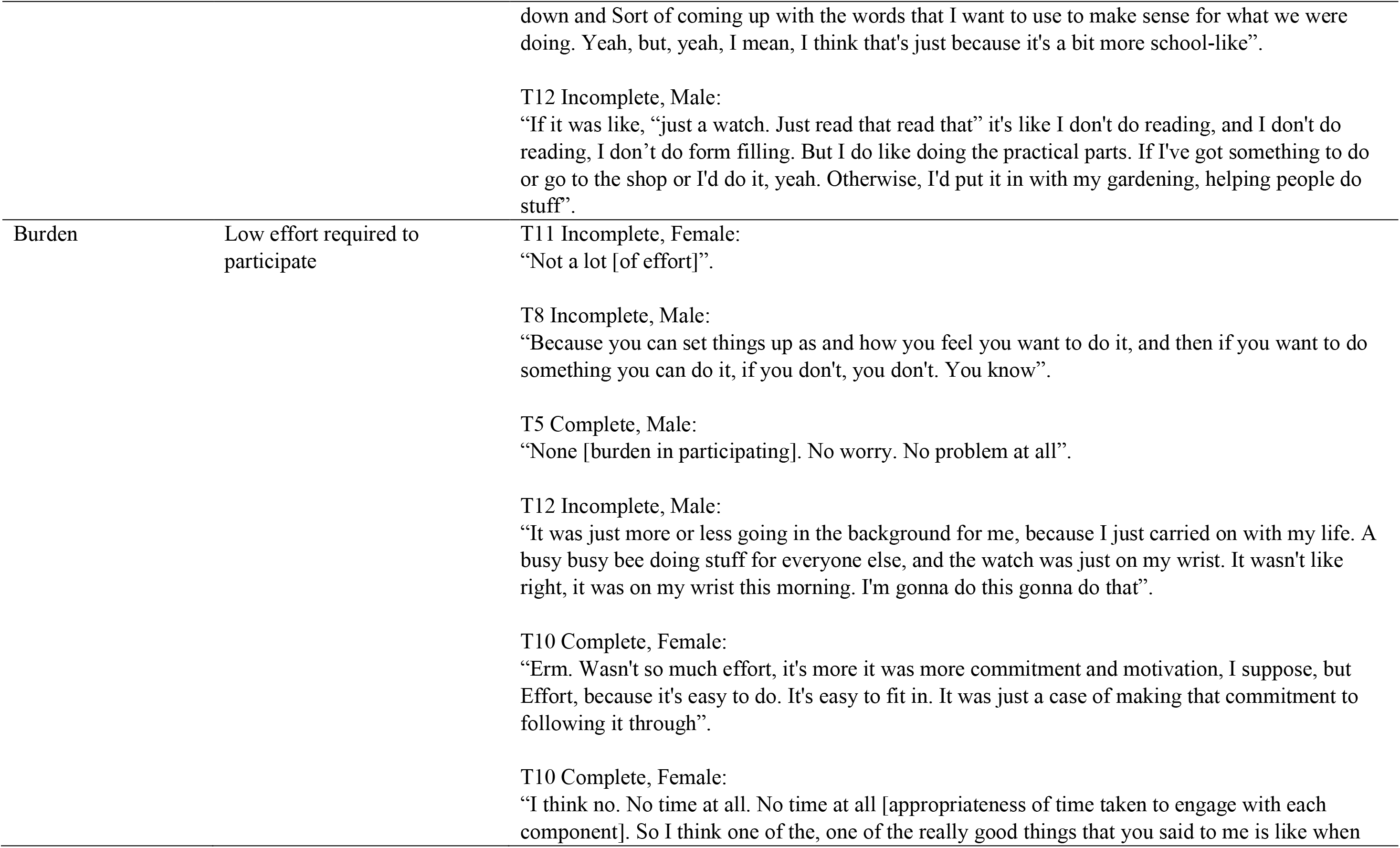
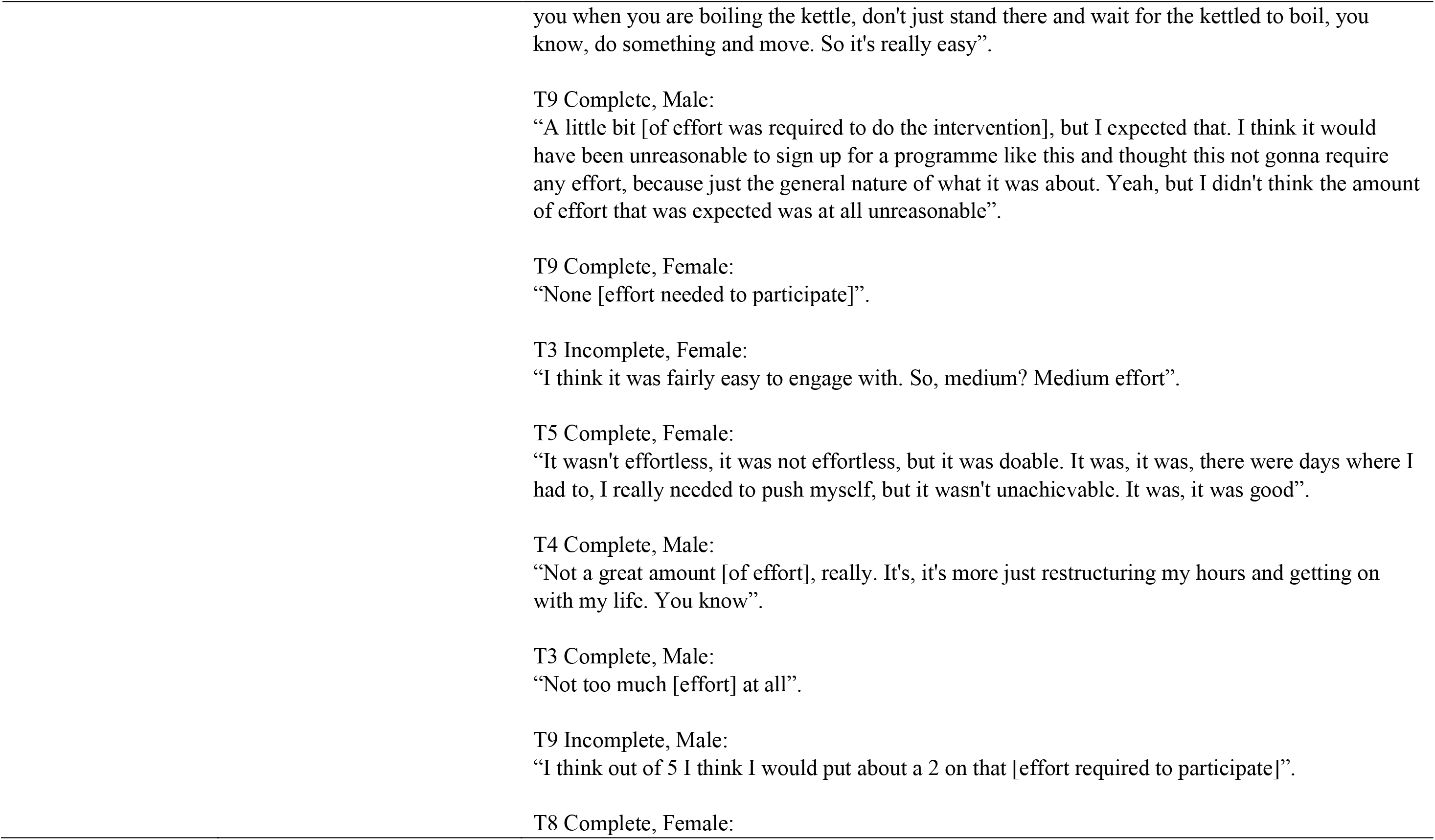
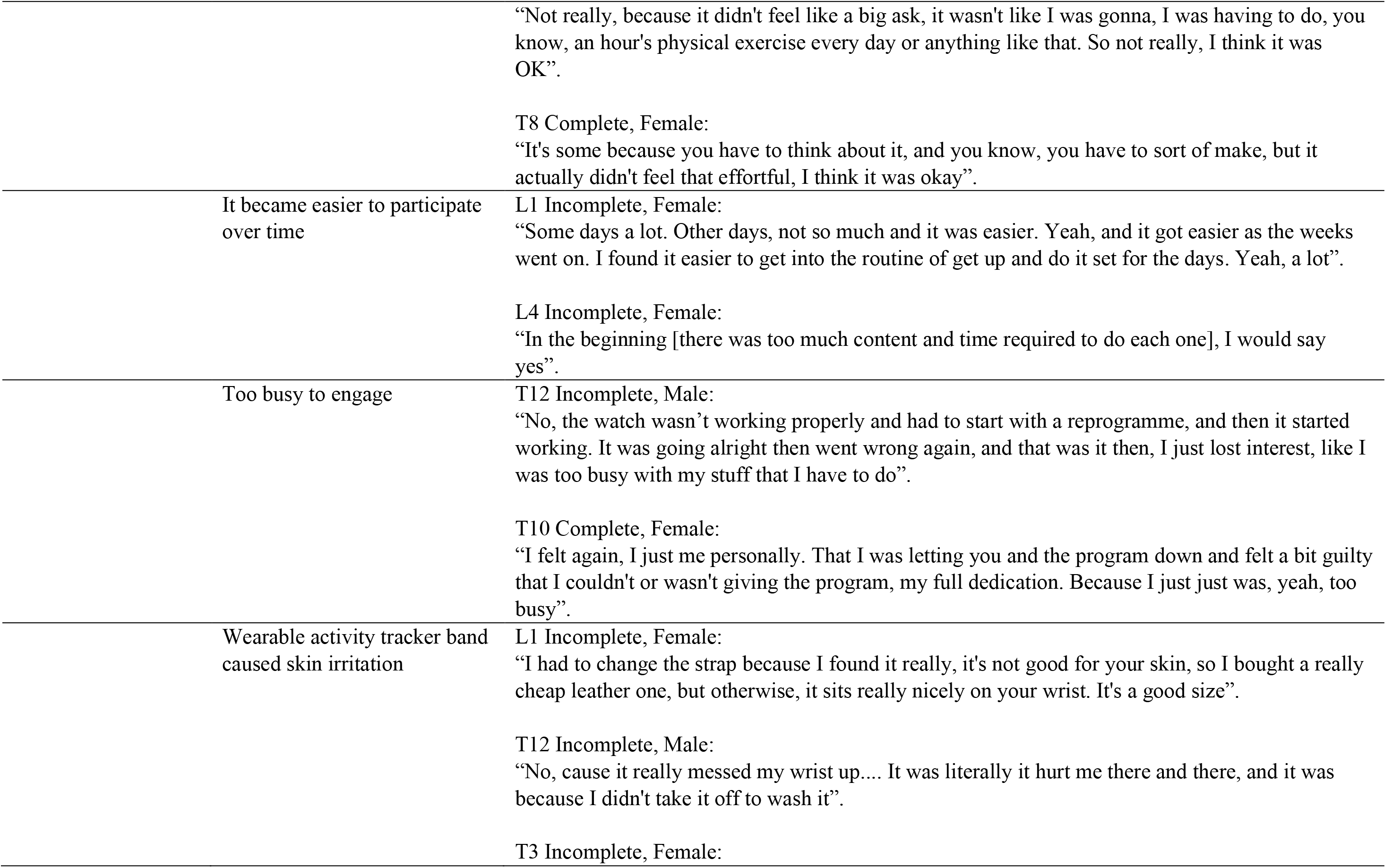
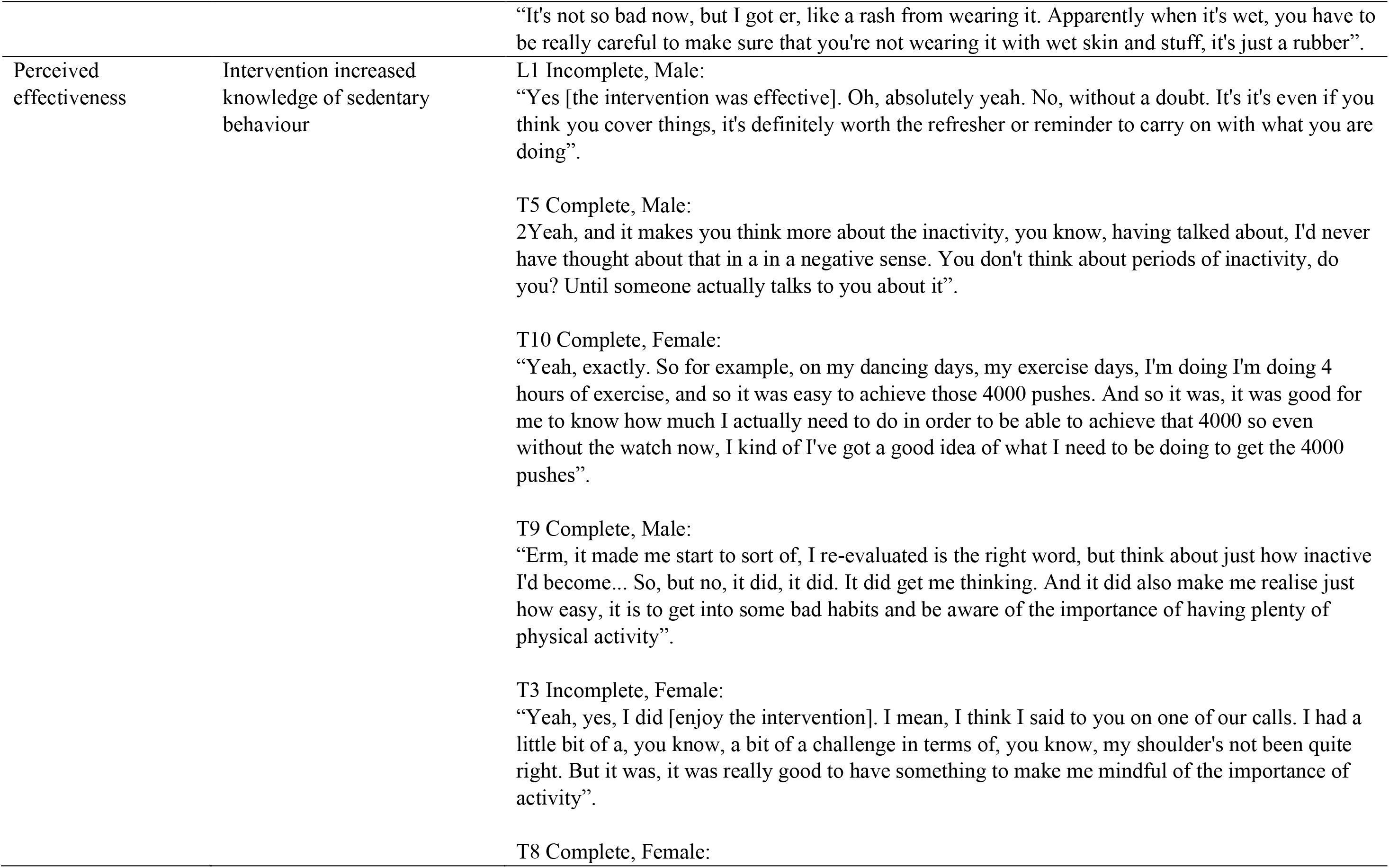
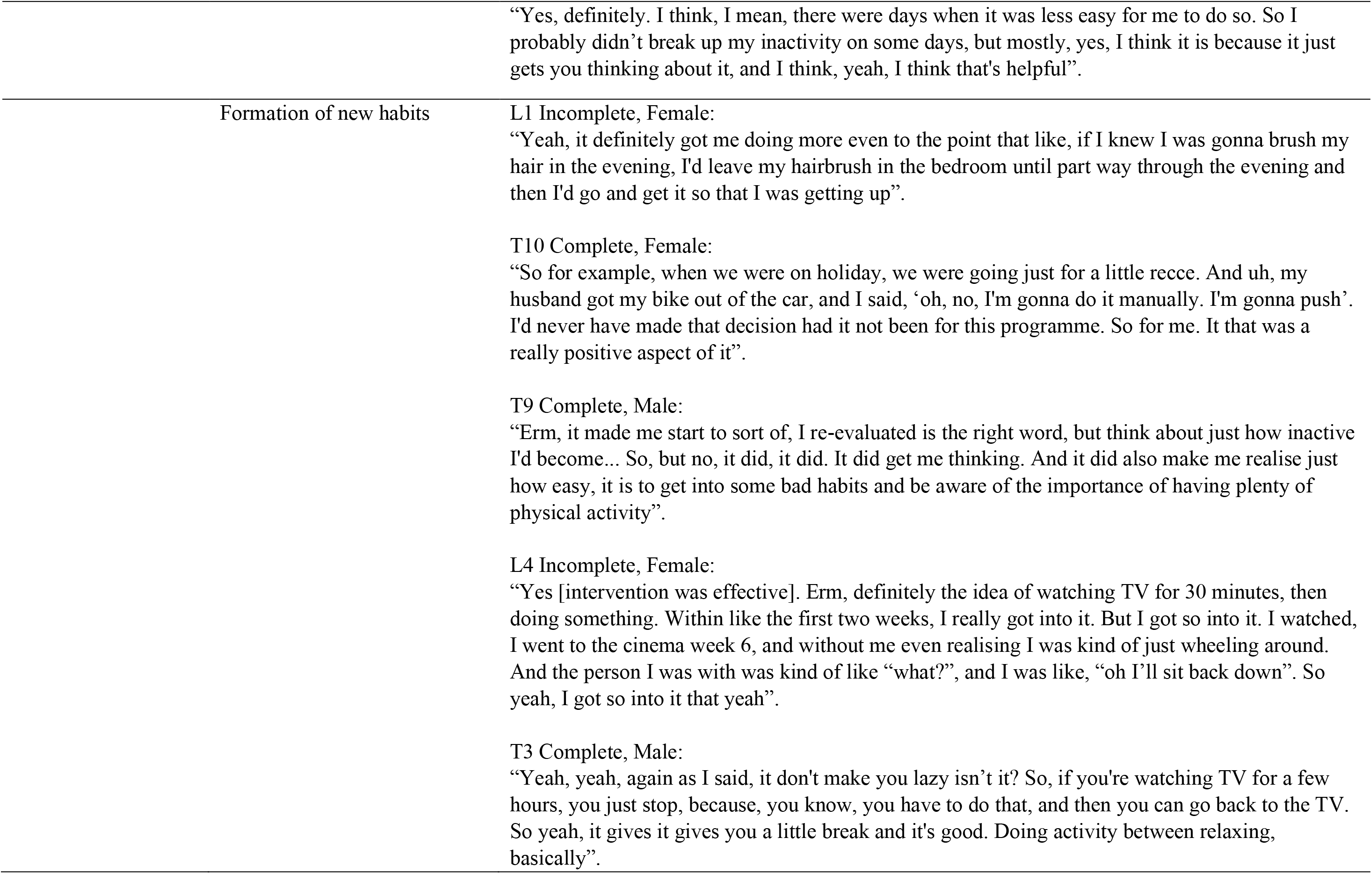
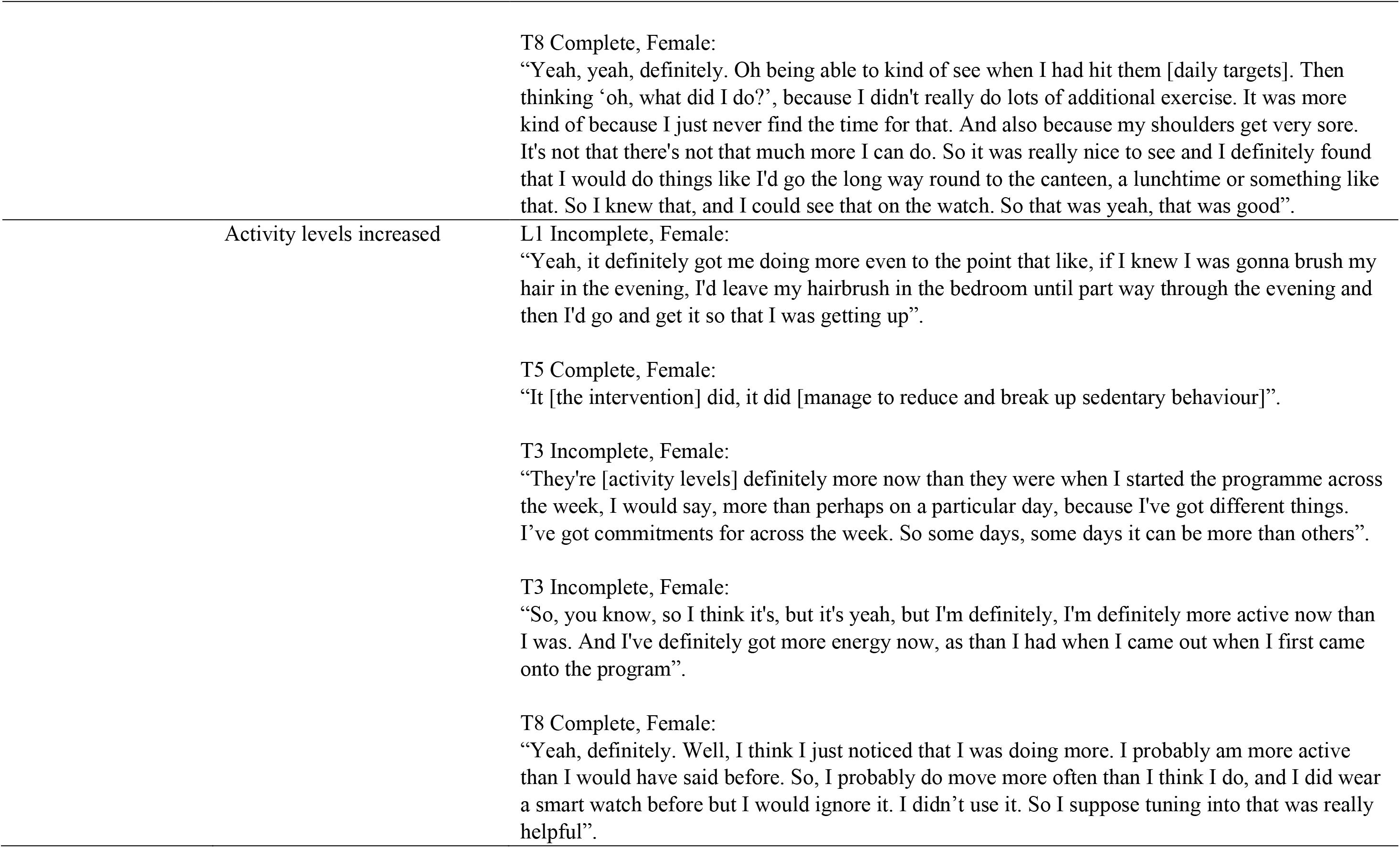
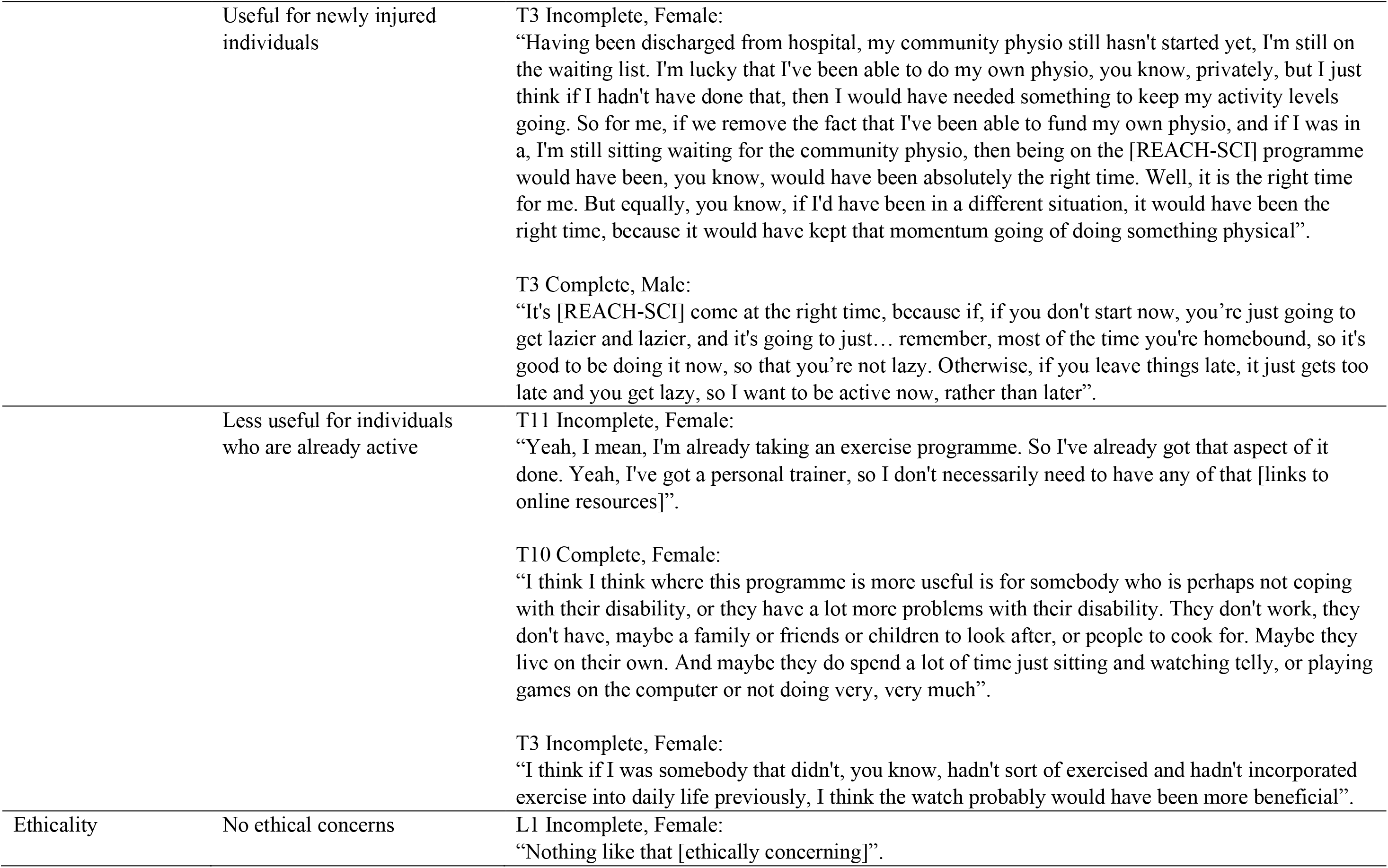
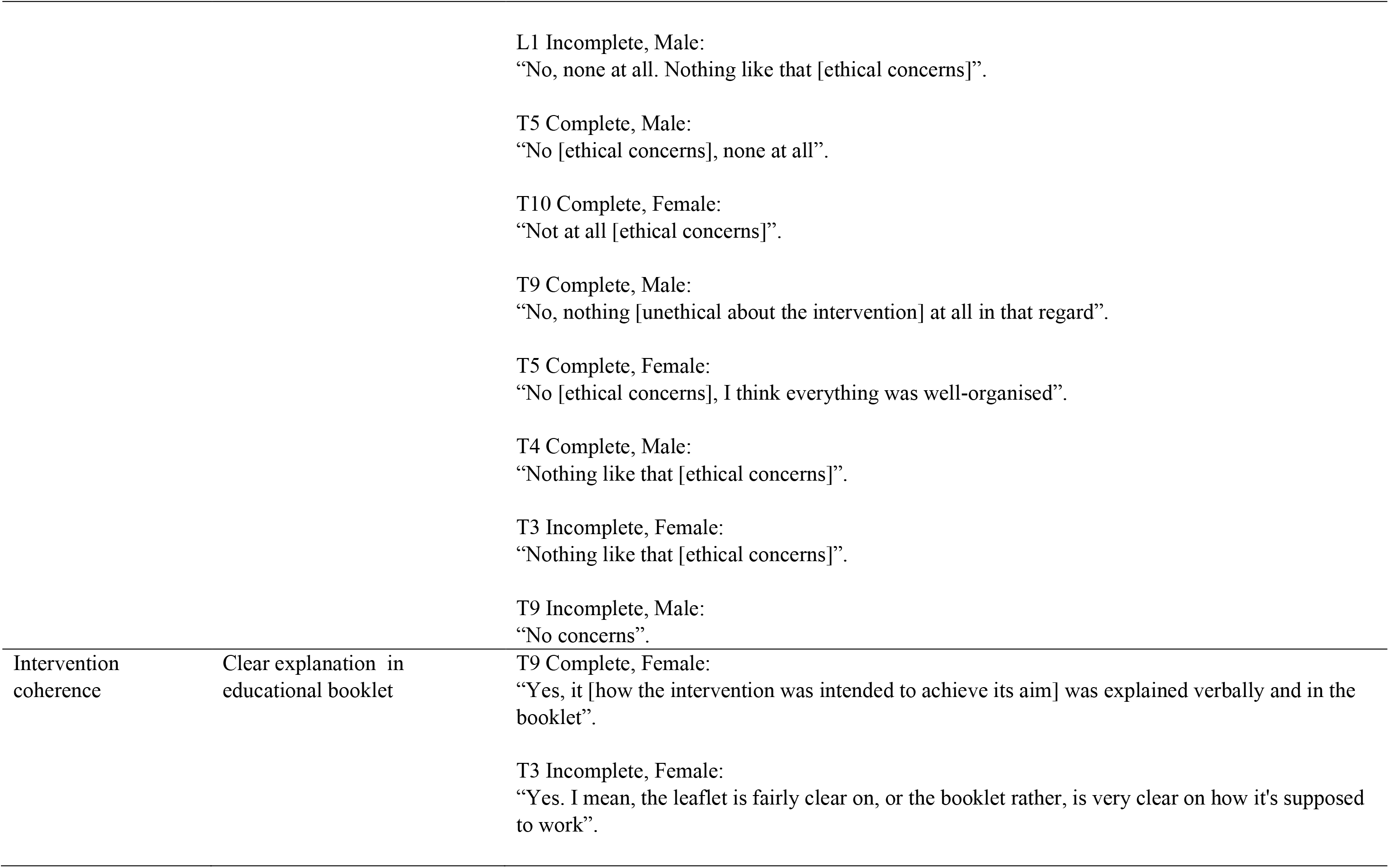
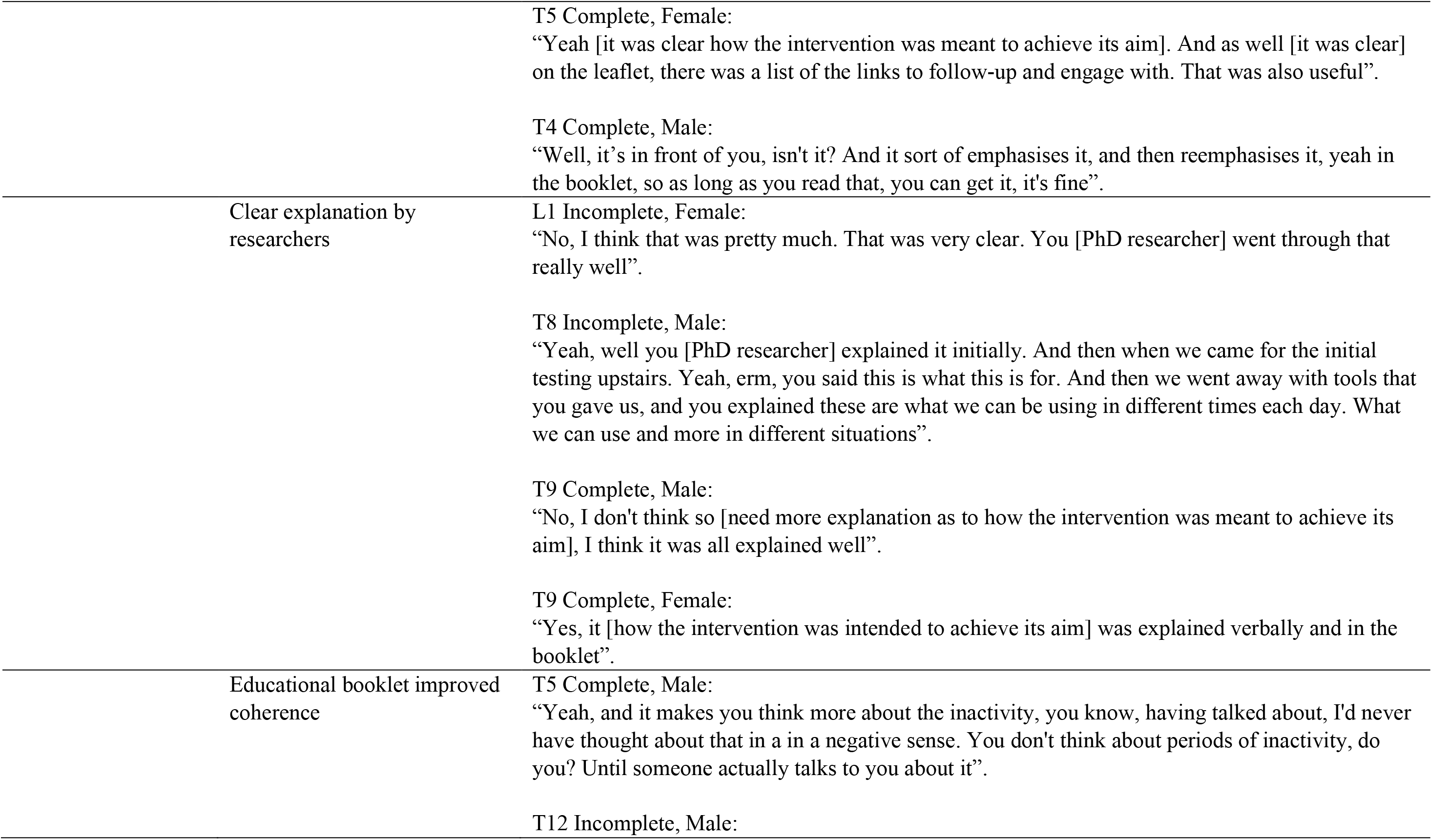
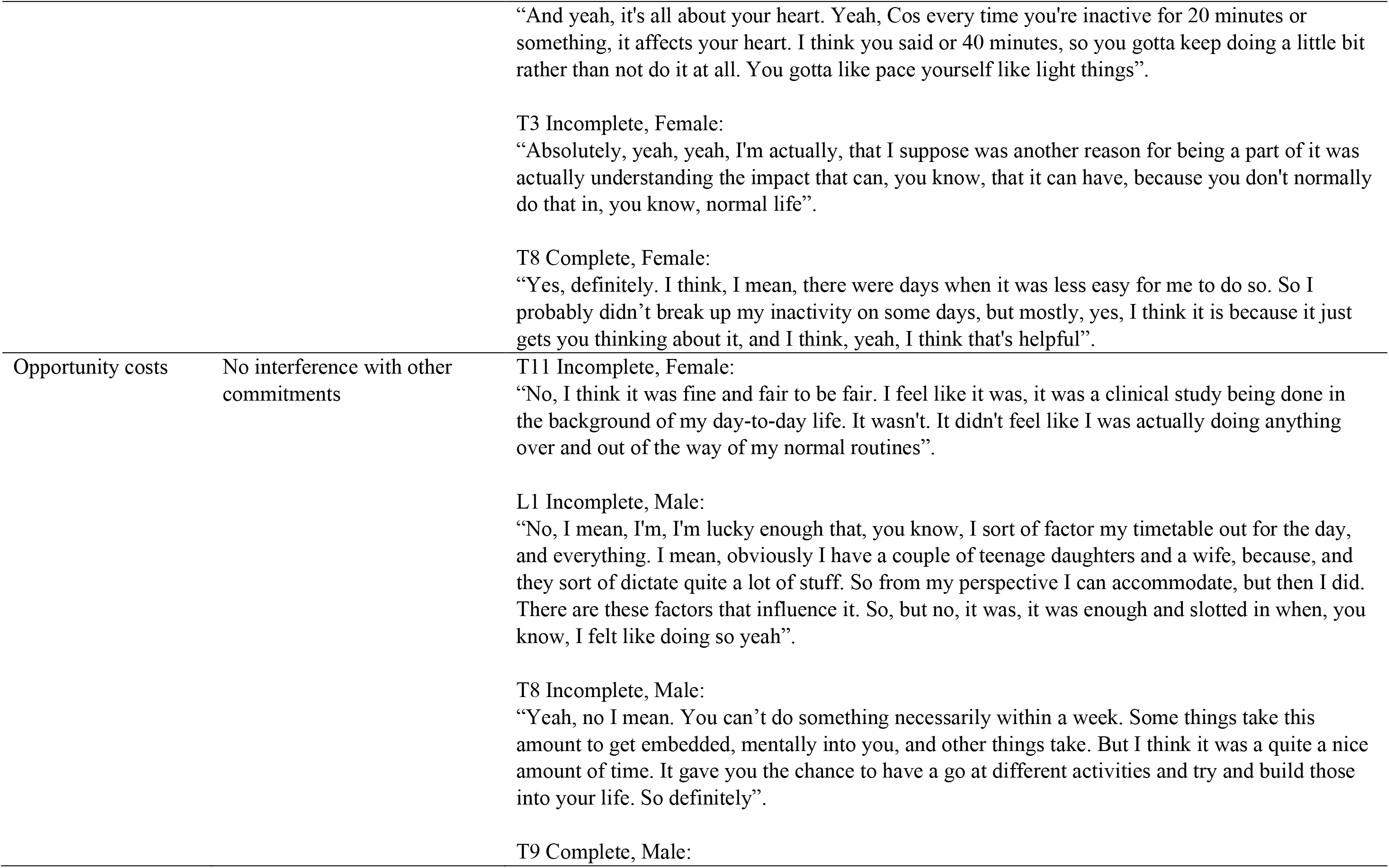
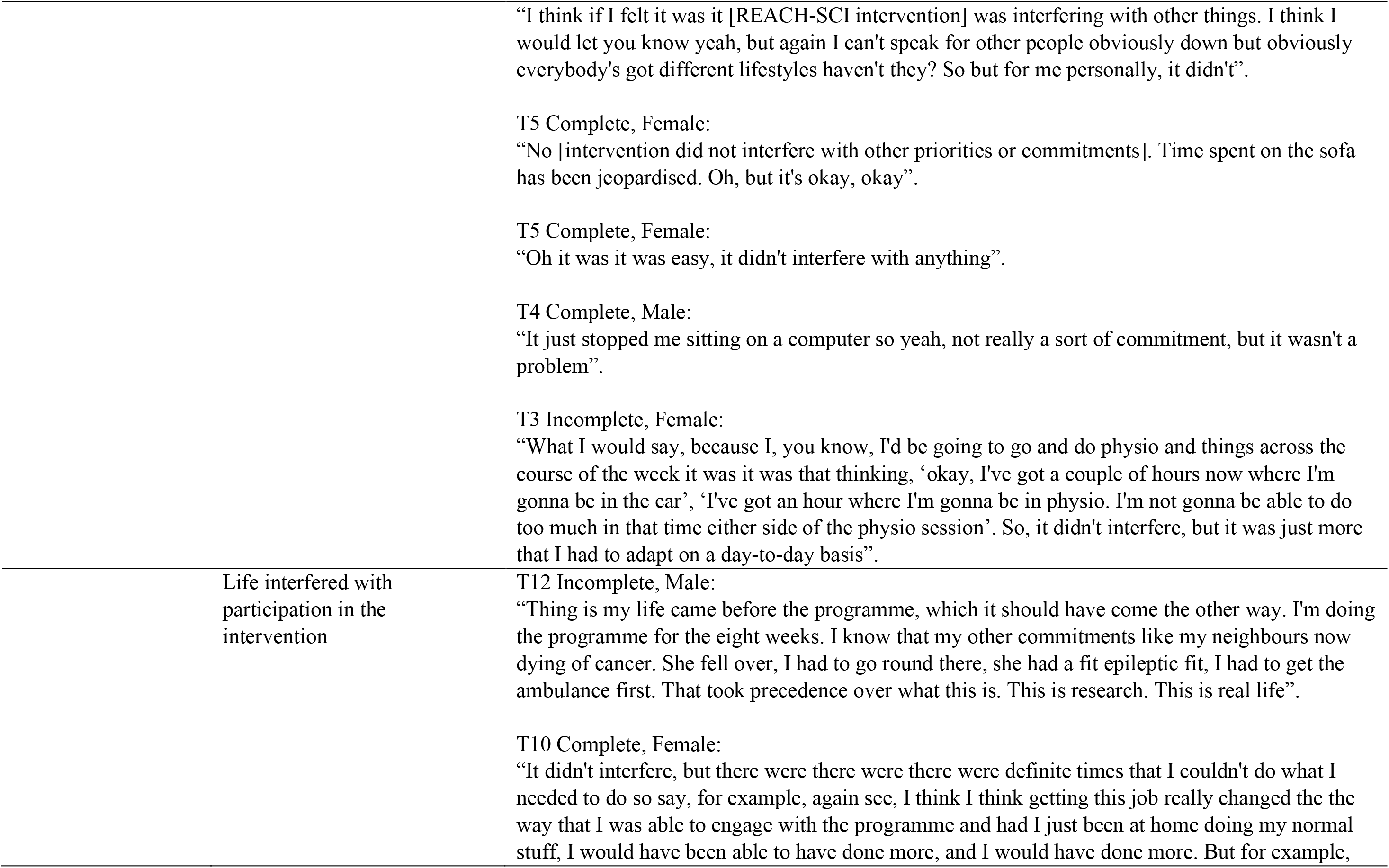
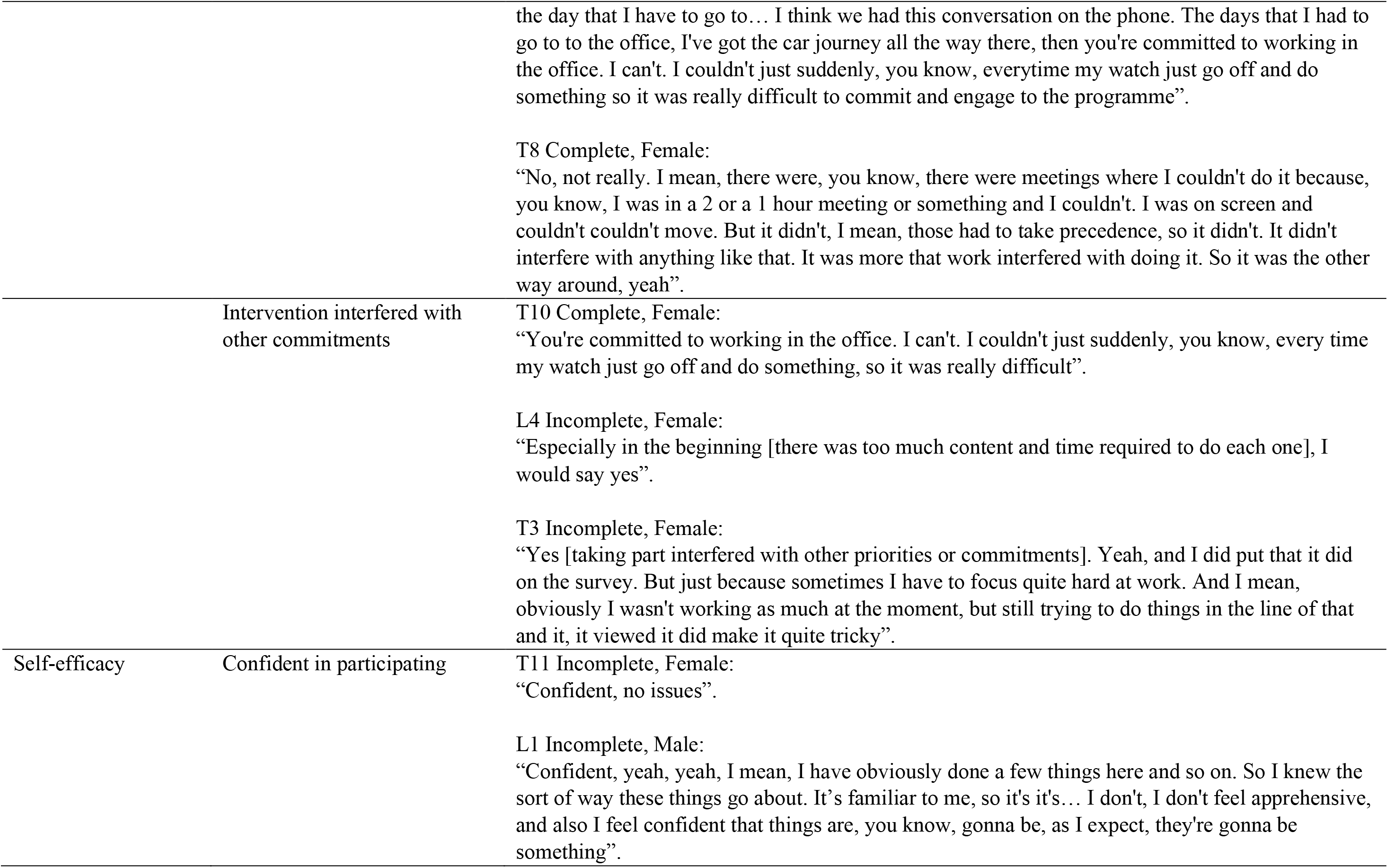
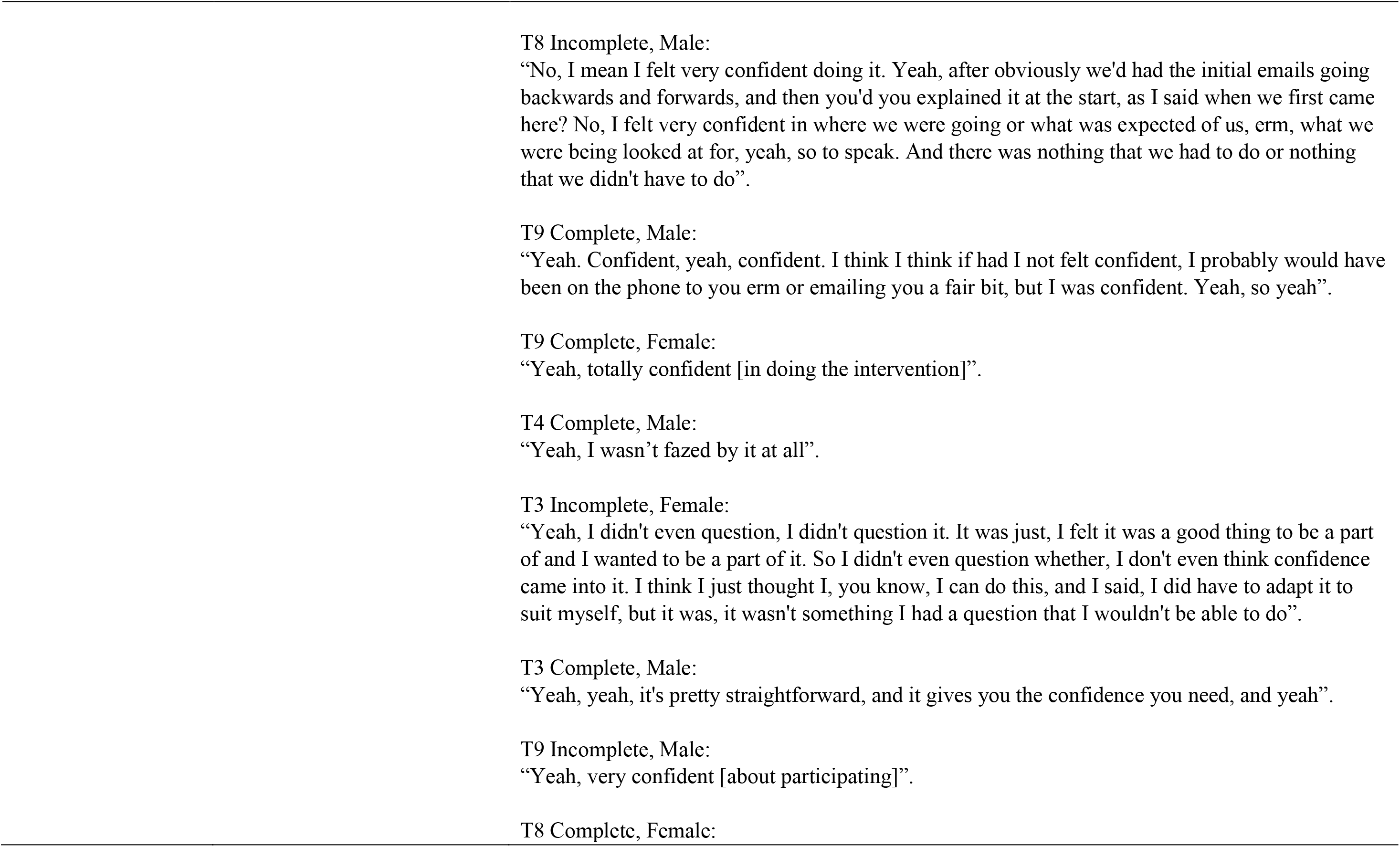
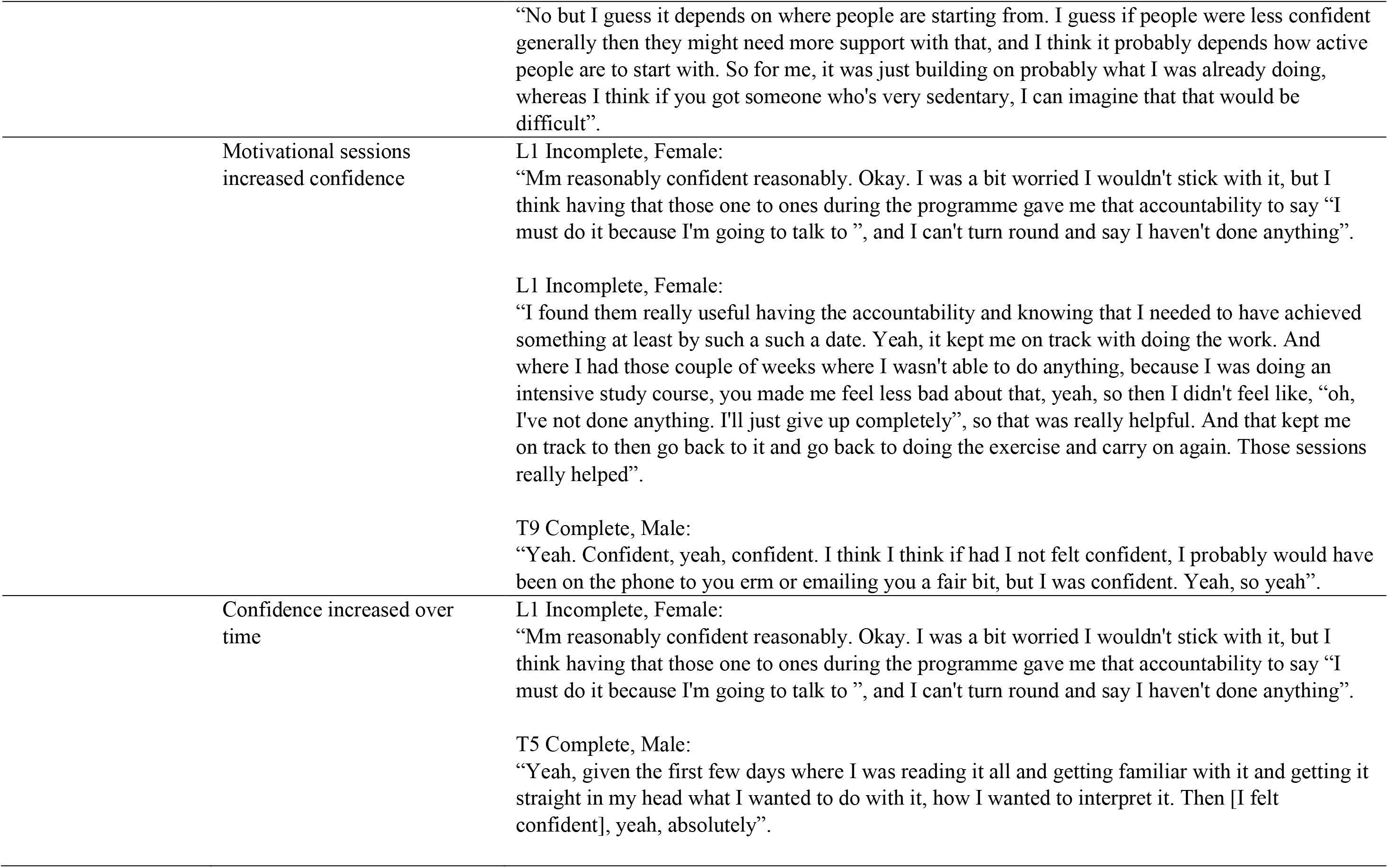
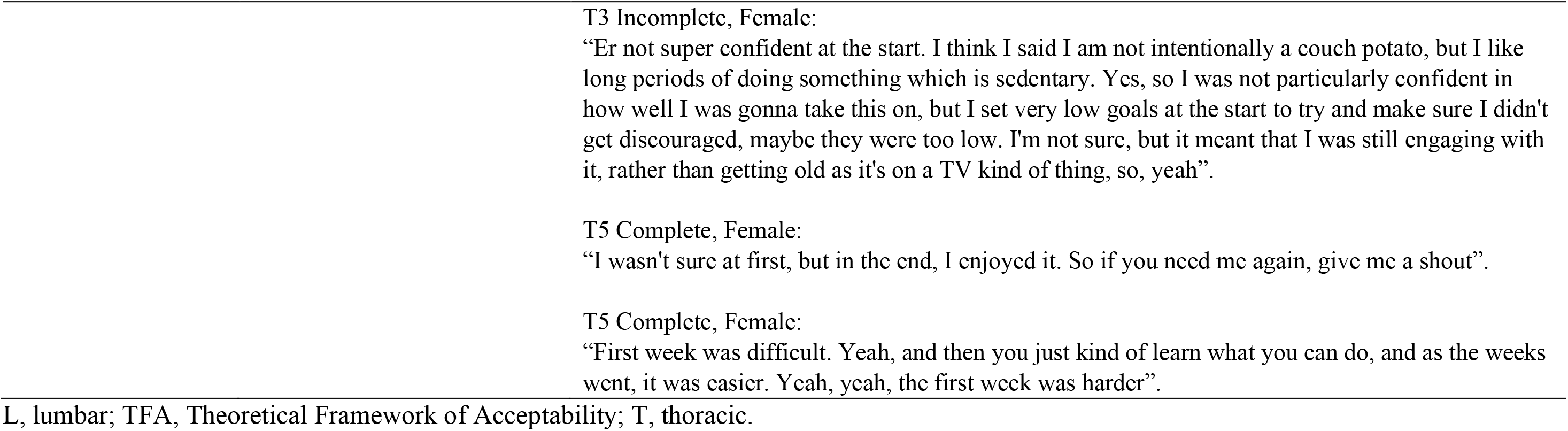
Intervention acceptability qualitative analysis with all quotations.

## Reference list

[1] Cirnigliaro CM, La Fountaine MF, Hobson JC, Kirshblum SC, Dengel DR, Spungen AM, et al. Predicting Cardiometabolic Risk From Visceral Abdominal Adiposity in Persons With Chronic Spinal Cord Injury. Journal of Clinical Densitometry 2021;24:442–52. 10.1016/j.jocd.2021.03.010.

[2] Gilbert O, Croffoot JR, Taylor AJ, Nash M, Schomer K, Groah S. Serum lipid concentrations among persons with spinal cord injury - A systematic review and meta-analysis of the literature. Atherosclerosis 2014;232:305–12. 10.1016/j.atherosclerosis.2013.11.028.

[3] Solinsky R, Betancourt L, Schmidt-Read M, Kupfer M, Owens M, Schwab JM, et al. Acute Spinal Cord Injury Is Associated With Prevalent Cardiometabolic Risk Factors. Arch Phys Med Rehabil 2022;103:696–701. 10.1016/j.apmr.2021.04.022.

[4] Gorgey AS, Dudley GA. Skeletal muscle atrophy and increased intramuscular fat after incomplete spinal cord injury. Spinal Cord 2007;45:304–9. 10.1038/sj.sc.3101968.

[5] Cragg JJ, Noonan VK, Krassioukov A, Borisoff J. Cardiovascular disease and spinal cord injury Results from a national population health survey. Neurology 2013;81:723–8. 10.1212/WNL.0b013e3182a1aa68.

[6] Tremblay MS, Aubert S, Barnes JD, Saunders TJ, Carson V, Latimer-Cheung AE, et al. Sedentary Behavior Research Network (SBRN) - Terminology Consensus Project process and outcome. International Journal of Behavioral Nutrition and Physical Activity 2017;14:75. 10.1186/s12966-017-0525-8.

[7] Wilmot EG, Edwardson CL, Achana FA, Davies MJ, Gorely T, Gray LJ, et al. Sedentary time in adults and the association with diabetes, cardiovascular disease and death: systematic review and meta-analysis. Diabetologia 2012;55:2895–905. 10.1007/S00125-012-2677-Z.

[8] Bailey DP, Hewson DJ, Champion RB, Sayegh SM. Sitting Time and Risk of Cardiovascular Disease and Diabetes: A Systematic Review and Meta-Analysis. Am J Prev Med 2019;57:408–16. 10.1016/J.AMEPRE.2019.04.015.

[9] Postma K, Bussmann JBJ, van Diemen T, Post MWM, Dekkers J, van Nes IJW, et al. Physical Activity and Sedentary Behavior From Discharge to 1 Year After Inpatient Rehabilitation in Ambulatory People With Spinal Cord Injury: A Longitudinal Cohort Study. Arch Phys Med Rehabil 2020;101:2061–70. 10.1016/j.apmr.2020.06.027.

[10] Ekelund U, Tarp J, Ding D, Sanchez-Lastra MA, Dalene KE, Anderssen SA, et al. Deaths potentially averted by small changes in physical activity and sedentary time: an individual participant data meta-analysis of prospective cohort studies. The Lancet 2026;407. 10.1016/S0140-6736(25)02219-6.

[11] Paddison S, Hexter B. Spinal Cord Injury. In: Lennon S, Ramdharry G, Verheyden G, editors. Physical Management for Neurological Conditions, vol. 1. 4th ed., London: Elsevier Science. London, UK; 2018, p. 171–203.

[12] Hadgraft NT, Brakenridge CL, Dunstan DW, Owen N, Healy GN, Lawler SP. Perceptions of the acceptability and feasibility of reducing occupational sitting: Review and thematic synthesis. International Journal of Behavioral Nutrition and Physical Activity 2018;15. 10.1186/s12966-018-0718-9.

[13] Aunger JA, Doody P, Greig CA. Interventions targeting sedentary behavior in non-working older adults: a systematic review. Maturitas 2018;116. 10.1016/j.maturitas.2018.08.002.

[14] De Cocker K, Veldeman C, De Bacquer D, Braeckman L, Owen N, Cardon G, et al. Acceptability and feasibility of potential intervention strategies for influencing sedentary time at work: Focus group interviews in executives and employees. International Journal of Behavioral Nutrition and Physical Activity 2015;12. 10.1186/s12966-015-0177-5.

[15] Cooper DL, Warland A, Norris E, Kilbride C, Paddison S, Bailey DP. Using the behaviour change wheel to co-design an intervention targeting sedentary behaviour in individuals with spinal cord injury. Clinical Rehabilitation 2026; 1–23. 10.1177/02692155261498670.

[16] van den Berg-Emons RJ, Bussmann JB, Haisma JA, Sluis TA, van der Woude LH, Bergen MP, et al. A Prospective Study on Physical Activity Levels After Spinal Cord Injury During Inpatient Rehabilitation and the Year After Discharge. Arch Phys Med Rehabil 2008;89:2094–101. 10.1016/j.apmr.2008.04.024.

[17] Cooper DL, Warland A, Norris E, Kilbride C, Paddison S, Bailey DP. Effects of interventions on sedentary behaviour and cardiovascular disease biomarkers in individuals with spinal cord injury: a systematic review. Disability and Rehabilitation 2026;48:4315–4338. 10.1080/09638288.2025.2592500.

[18] Eldridge SM, Chan CL, Campbell MJ, Bond CM, Hopewell S, Thabane L, et al. CONSORT 2010 statement: Extension to randomised pilot and feasibility trials. The BMJ 2016;355:i5239. 10.1136/bmj.i5239.

[19] O’Cathain A, Murphy E, Nicholl J. The quality of mixed methods studies in health services research. J Health Serv Res Policy 2008;13:92–8. 10.1258/jhsrp.2007.007074.

[20] Warburton D, Jamnik V, Bredin S, Shephard R,, Gledhill N. The 2021 physical activity readiness questionnaire for everyone (PAR-Q+) and electronic physical activity readiness medical examination (ePARmed-X+): 2021 PAR-Q+. The Health & Fitness Journal of Canada 2021;14:83–7. 10.14288/hfjc.v14i1.351.

[21] Teresi JA, Yu X, Stewart AL, Hays RD. Guidelines for Designing and Evaluating Feasibility Pilot Studies. Med Care 2022;60:95–103. 10.1097/MLR.0000000000001664.

[22] Skivington K, Matthews L, Simpson SA, Craig P, Baird J, Blazeby JM, et al. A new framework for developing and evaluating complex interventions: Update of Medical Research Council guidance. The BMJ 2021;374:n2061. 10.1136/bmj.n2061.

[23] Michie S, van Stralen MM, West R. The behaviour change wheel: A new method for characterising and designing behaviour change interventions. Implementation Science 2011;6:42. 10.1186/1748-5908-6-42.

[24] Hoffmann TC, Glasziou PP, Boutron I, Milne R, Perera R, Moher D, et al. Better reporting of interventions: Template for intervention description and replication (TIDieR) checklist and guide. BMJ (Online) 2014;348:g1687. 10.1136/bmj.g1687.

[25] Sekhon M, Cartwright M, Francis JJ. Development of a theory-informed questionnaire to assess the acceptability of healthcare interventions. BMC Health Serv Res 2022;22:279. 10.1186/s12913-022-07577-3.

[26] Sekhon M, Cartwright M, Francis JJ. Acceptability of healthcare interventions: An overview of reviews and development of a theoretical framework. BMC Health Serv Res 2017;17:88. 10.1186/s12913-017-2031-8.

[27] Melzack R. The McGill Pain Questionnaire: Major properties and scoring methods. Pain 1975;1:277–99. 10.1016/0304-3959(75)90044-5.

[28] Fisk JD, Ritvo PG, Ross L, Haase DA, Marrie TJ, Schlech WF. Measuring the functional impact of fatigue: Initial validation of the fatigue impact scale. Clinical Infectious Diseases 1994;18:s79–83. 10.1093/clinids/18.Supplement_1.S79.

[29] Imam B, Anton HA, Miller WC. Measurement properties of a telephone version of the Modified Fatigue Impact Scale among individuals with a traumatic spinal cord injury. Spinal Cord 2012;50:920–4. 10.1038/sc.2012.79.

[30] Sawatzky B, Bishop CM, Miller WC. Classification and measurement of pain in the spinal cord-injured population. Spinal Cord 2008;46:2–10. 10.1038/sj.sc.3102137.

[31] Migueles JH, Rowlands A V, Huber F, Sabia S, Van Hees VT. GGIR: A Research Community–Driven Open Source R Package for Generating Physical Activity and Sleep Outcomes From Multi-Day Raw Accelerometer Data. J Meas Phys Behav 2019;2. 10.1123/jmpb.2018-0063.

[32] Migueles JH, Cadenas-Sanchez C, Ekelund U, Delisle Nyström C, Mora-Gonzalez J, Löf M, et al. Accelerometer Data Collection and Processing Criteria to Assess Physical Activity and Other Outcomes: A Systematic Review and Practical Considerations. Sports Medicine 2017;47:1821–45. 10.1007/s40279-017-0716-0.

[33] Van Hees VT, Fang Z, Langford J, Assah F, Mohammad A, Da Silva ICM, et al. Autocalibration of accelerometer data for free-living physical activity assessment using local gravity and temperature: An evaluation on four continents. J Appl Physiol 2014;117:738–44. 10.1152/japplphysiol.00421.2014.

[34] Bailey DP, Ahmed I, Cooper DL, Finlay KA, Froome HM, Nightingale TE, et al. Validity of a wrist-worn consumer-grade wearable for estimating energy expenditure, sedentary behaviour, and physical activity in manual wheelchair users with spinal cord injury. Disabil Rehabil Assist Technol 2025;20:708–14. 10.1080/17483107.2024.2405895.

[35] Office for National Statistics. Analysis of Experimental Subjective Wellbeing Data from the Annual Population Survey 2011. https://data.parliament.uk/DepositedPapers/Files/DEP2013-1761/95FC57F0.pdf (accessed May 9, 2026).

[36] Spitzer RL, Kroenke K, Williams JBW, Löwe B. A brief measure for assessing generalized anxiety disorder: The GAD-7. Arch Intern Med 2006;166:1092–7. 10.1001/archinte.166.10.1092.

[37] Kroenke K, Spitzer RL, Williams JBW. The PHQ-9: Validity of a brief depression severity measure. J Gen Intern Med 2001;16:606–13. 10.1046/j.1525-1497.2001.016009606.x.

[38] Ware JE, Sherbourne CD. The MOS 36-item short-form health survey (Sf-36): I. conceptual framework and item selection. Med Care 1992;30:473–83. 10.1097/00005650-199206000-00002.

[39] Lee BB, Simpson JM, King MT, Haran MJ, Marial O. The SF-36 walk-wheel: A simple modification of the SF-36 physical domain improves its responsiveness for measuring health status change in spinal cord injury. Spinal Cord 2009;47:50–5. 10.1038/sc.2008.65.

[40] Gale NK, Heath G, Cameron E, Rashid S, Redwood S. Using the framework method for the analysis of qualitative data in multi-disciplinary health research. BMC Med Res Methodol 2013;13:117. 10.1186/1471-2288-13-117.

[41] Creswell JW. A concise introduction to mixed methods research. Sage Publishing. Thousand Oaks, California; 2021.

[42] Dogra S, Copeland JL, Altenburg TM, Heyland DK, Owen N, Dunstan DW. Start with reducing sedentary behavior: A stepwise approach to physical activity counseling in clinical practice. Patient Educ Couns 2022;105:1353–61. 10.1016/j.pec.2021.09.019.

[43] Ezeugwu VE, Manns PJ. The Feasibility and Longitudinal Effects of a Home-Based Sedentary Behavior Change Intervention After Stroke. Arch Phys Med Rehabil 2018;99:2540–7. 10.1016/j.apmr.2018.06.014.

[44] Manns PJ, Mehrabani G, Norton S, Aminian S, Motl RW. The SitLess With MS Program: Intervention Feasibility and Change in Sedentary Behavior. Arch Rehabil Res Clin Transl 2020;2:100083. 10.1016/j.arrct.2020.100083.

[45] English C, Healy GN, Olds T, Parfitt G, Borkoles E, Coates A, et al. Reducing Sitting Time After Stroke: A Phase II Safety and Feasibility Randomized Controlled Trial. Arch Phys Med Rehabil 2016;97:273–80. 10.1016/j.apmr.2015.10.094.

[46] Dyson-Hudson TA, Kirshblum SC. Shoulder Pain In Chronic Spinal Cord Injury, Part 1: Epidemiology, Etiology, And Pathomechanics. J Spinal Cord Med 2004;27:4–17. 10.1080/10790268.2004.11753724.

[47] McGowan LJ, Chater AM, Harper JH, Kilbride C, Victor C, Brierley ML, et al. Acceptability of a remotely delivered sedentary behaviour intervention to improve sarcopenia and maintain independent living in older adults with frailty: a mixed-methods study. BMC Geriatr 2024;24:820. 10.1186/s12877-024-05385-4.

[48] Carr LJ, Karvinen K, Peavler M, Smith R, Cangelosi K. Multicomponent intervention to reduce daily sedentary time: A randomised controlled trial. BMJ Open 2013;3:e003261. 10.1136/bmjopen-2013-003261.

[49] Nieste I, Franssen WMA, Spaas J, Bruckers L, Savelberg HHCM, Eijnde BO. Lifestyle interventions to reduce sedentary behaviour in clinical populations: A systematic review and meta-analysis of different strategies and effects on cardiometabolic health. Prev Med (Baltim) 2021;148:106593. 10.1016/j.ypmed.2021.106593.

[50] Smith-Turchyn J, Sinclair S, O’Loughlin EK, Innes A, Vani MF, Beauchamp M, et al. A pilot randomized controlled trial of a virtual peer-support exercise intervention for female older adults with cancer. BMC Geriatr 2024;24:887. 10.1186/s12877-024-05495-z.

[51] Brierley ML, Chater AM, Edwardson CL, Castle EM, Hunt ER, Biddle SJH, et al. The Regulate your Sitting Time (RESIT) intervention for reducing sitting time in individuals with type 2 diabetes: findings from a randomised-controlled feasibility trial. Diabetology and Metabolic Syndrome 2024;16:87. 10.1186/s13098-024-01336-6.

[52] Martinez JM, Haubert LL, Eberly VJ, Weiss WB, Rankin JW. A randomized comparative effectiveness trial to evaluate two programs for promotion of physical activity after spinal cord injury in manual wheelchair users. Front Sports Act Living 2025;7:1504840. 10.3389/FSPOR.2025.1504840/BIBTEX.

[53] Freire YA, Browne RAV, Cabral LLP, Barreira T V., Costa EC. Moderating effects of physical activity intensity and sedentary break patterns on the association between sedentary time and cardiometabolic risk in older adults. Physiol Behav 2026;306. 10.1016/j.physbeh.2025.115214.

[54] Katzmarzyk PT, Ross R, Blair SN, Després JP. Should we target increased physical activity or less sedentary behavior in the battle against cardiovascular disease risk development? Atherosclerosis 2020;311:107–15. 10.1016/j.atherosclerosis.2020.07.010.

[55] Després JP. Body fat distribution and risk of cardiovascular disease: An update. Circulation 2012;126:1301–13. 10.1161/CIRCULATIONAHA.111.067264.

[56] NHS. High cholesterol - Cholesterol levels - NHS 2026. https://www.nhs.uk/conditions/high-cholesterol/cholesterol-levels/ (accessed April 29, 2026).

[57] Mehrabani G, Aminian S, Norton S, Motl RW, Manns PJ. Preliminary efficacy of the “SitLess with MS” intervention for changing sedentary behaviour, symptoms, and physical performance in multiple sclerosis. Disabil Rehabil 2022;44:6374–81. 10.1080/09638288.2021.1966520.

[58] Bresnahan JJ, Scoblionko BR, Zorn D, Graves DE, Viscusi ER. The demographics of pain after spinal cord injury: a survey of our model system. Spinal Cord Ser Cases 2022;8. 10.1038/s41394-022-00482-1.

